# Machine Learning and Explainable AI for Multi-State Classification of Malaria Transmission Dynamics in Kenya

**DOI:** 10.64898/2026.05.09.26352789

**Authors:** Maurice Wanyonyi, Jacqueline Akelo Gogo

## Abstract

Malaria remains a major public health challenge in sub–Saharan Africa, with pronounced spatial and temporal variation in transmission intensity that complicates effective control strategies. Accurate classification of transmission states is essential for guiding targeted interventions and strengthening early warning systems. This study develops a machine learning framework for the classification of malaria transmission states in Kenya using monthly panel data from 47 counties spanning the period 2015 to 2025. Transmission was categorised into four operationally relevant states based on incidence thresholds. Four supervised learning models, namely multinomial logistic regression, random forest, extreme gradient boosting, and support vector machine, were trained using temporally lagged features and evaluated under a forward chaining validation scheme to preserve temporal structure. Model performance was assessed using accuracy, macro averaged F1 score, Matthews correlation coefficient, and Brier score, complemented by calibration analysis. Extreme gradient boosting achieved the best overall performance, with accuracy of 0.9918, macro averaged F1 score of 0.9647, and Matthews correlation coefficient of 0.9831, alongside the lowest Brier score of 0.0031, indicating highly reliable probability estimates. Feature importance analysis revealed that lagged incidence, vegetation index, precipitation, and insecticide treated net coverage were the most influential predictors. Partial dependence analysis demonstrated nonlinear relationships and clear seasonal patterns in transmission dynamics. The findings show that machine learning approaches can accurately classify malaria transmission states while providing interpretable and well calibrated outputs for decision making. This framework offers a practical tool for supporting malaria surveillance and resource allocation. Further validation in different epidemiological settings is recommended to assess generalisability.

## Introduction

Malaria remains a major public health burden in sub Saharan Africa, with an estimated 249 million cases and 608000 deaths worldwide in 2022, of which 94% of cases and 95% of deaths occurred in the African Region [1]. In Kenya, malaria transmission exhibits substantial spatial and temporal heterogeneity, ranging from highly endemic regions around Lake Victoria and coastal zones to epidemic prone highland areas and arid regions with low transmission [2, 3]. Approximately 70% of the population is at risk, and malaria accounts for over 20% of outpatient visits in high burden settings [4]. Despite sustained scale up of insecticide treated nets, indoor residual spraying, and artemisinin based combination therapies, progress in malaria control has slowed, with recent evidence of resurgence linked to insecticide resistance, climate variability, and declining intervention effectiveness [5, 6]. These challenges highlight the need for improved analytical approaches that can support timely and targeted intervention strategies.

Accurate characterisation and prediction of malaria transmission dynamics are essential for guiding resource allocation, informing policy decisions, and strengthening early warning systems. Traditional modelling approaches, including compartmental models and statistical time series methods, have provided important insights into malaria dynamics [7, 8]. However, these approaches often rely on restrictive assumptions such as linear relationships, stationarity, and predefined functional forms, which limit their ability to represent the complex and nonlinear interactions among climatic, environmental, and intervention related factors. In addition, their capacity to incorporate high dimensional data and capture context specific variability across space and time remains limited.

Machine learning methods offer a flexible and data driven alternative by learning patterns directly from observed data without requiring explicit specification of underlying relationships [9, 10]. These approaches are well suited to modelling complex epidemiological systems, as they can capture nonlinear dependencies, represent interactions among multiple predictors, and improve predictive accuracy in settings characterised by heterogeneity and uncertainty. Consequently, machine learning has gained increasing attention in malaria research.

Several studies have demonstrated the potential of machine learning in this domain. Ayoka and Nnadi [11] applied machine learning techniques to predict malaria prevalence in Nigeria, while Baba Adamu et al. [12] examined the influence of climate variability on malaria transmission. Morang’a et al. [13] used machine learning to classify clinical malaria outcomes based on haematological indicators. Taconet et al. [14] developed interpretable models to analyse environmental drivers of mosquito biting rates in Burkina Faso, and Zheng et al. [15] investigated climate driven transmission patterns in Tanzania. Additional work has explored integration with spatial clustering and mathematical modelling frameworks [16, 17]. While these studies provide valuable insights, most focus on continuous outcomes such as incidence or prevalence and often do not directly align with operational decision making frameworks used in malaria control.

Despite recent progress, several important gaps remain. First, limited attention has been given to modelling malaria transmission as discrete operational states that correspond to actionable risk categories used in public health practice. Second, many existing studies emphasise predictive accuracy without adequately assessing probabilistic calibration, even though reliable probability estimates are critical for risk based decision making. Third, the application of explainable artificial intelligence methods remains limited, reducing transparency and hindering practical adoption of machine learning models in public health settings. Fourth, computational considerations such as training time, scalability, and inference efficiency are rarely evaluated, despite their importance for implementation in resource constrained environments.

To address these gaps, this study develops a comprehensive and interpretable machine learning framework for the classification of malaria transmission states in Kenya using spatio temporal panel data. The study models transmission as discrete epidemiologically meaningful categories, evaluates multiple machine learning algorithms using both predictive and calibration metrics, and integrates explainable artificial intelligence methods to enhance interpretability. In addition, computational performance is assessed to ensure that the proposed framework is suitable for practical deployment in real world settings.

The contributions of this study are fourfold. First, it introduces a multi state classification framework that directly predicts operationally relevant transmission categories. Second, it provides a rigorous comparative evaluation of machine learning models using both accuracy based and probabilistic calibration measures. Third, it integrates explainable artificial intelligence techniques to identify key drivers of malaria transmission and improve model transparency. Fourth, it evaluates computational efficiency to support implementation in routine surveillance systems.

Overall, this study presents a robust, interpretable, and scalable framework for modelling malaria transmission dynamics in Kenya, with direct relevance for evidence based malaria control and early warning systems.

## Materials and methods

### Study Design and Data Sources

This study adopts a quantitative longitudinal design using a panel dataset comprising monthly observations for all 47 counties in Kenya from January 2015 to December 2025. The unit of analysis is the county month, resulting in a balanced panel with complete temporal coverage across all spatial units.

Malaria incidence data were obtained from the Kenya Ministry of Health District Health Information System version two and supplemented with Kenya Malaria Indicator Surveys conducted in 2015, 2020, and 2025. Environmental variables, including temperature, precipitation, and Normalised Difference Vegetation Index, were obtained from the Climate Hazards Group InfraRed Precipitation with Station data. Intervention related variables, including insecticide treated net coverage, were derived from Malaria Indicator Surveys and routine distribution records. Static geographical variables, including elevation and population density, were obtained from the Shuttle Radar Topography Mission and the Kenya National Bureau of Statistics.

All datasets were harmonised to a common monthly temporal resolution and aligned spatially at the county level using consistent administrative identifiers.

The outcome variable is a categorical malaria transmission state denoted by *S*_*i,t*_ ∈ {0, 1, 2, 3}, where *i* = 1, …, 47 indexes counties and *t* indexes time in months. The transmission state is derived from malaria incidence per 1000 population, denoted by *I*_*i,t*_, as follows:

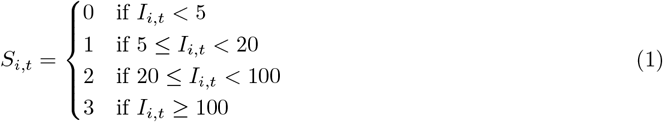

These thresholds are consistent with epidemiological stratification approaches used in malaria surveillance and reflect operational categories relevant for intervention planning [2, 3].

### Data Preprocessing

All preprocessing steps were implemented within a training data pipeline to avoid information leakage.

Continuous variables were standardised using statistics computed from the training data only and subsequently applied to validation and test sets.

Missing values in environmental variables were imputed using linear interpolation for short gaps and seasonal mean imputation for longer gaps. Intervention and demographic variables were imputed using last observation carried forward when appropriate to preserve temporal consistency.

Outliers were assessed using interquartile range criteria. Observations were retained unless clear data entry errors were identified, in which case corrections were applied based on source verification. All preprocessing procedures were implemented using Python libraries including pandas and numpy.

### Feature Engineering and Temporal Structure

Let *X*_*i,t*_ ∈ R^*p*^ denote the vector of observed covariates for county *i* at time *t*. To capture temporal dependence and delayed effects, lagged features were constructed.

For lag order *l* ∈ {1, 2}, lagged covariates are defined as:

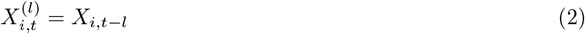

Lagged outcome states were included to represent persistence in transmission dynamics:

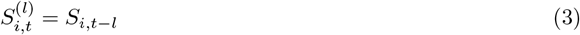

The final feature vector is defined as:

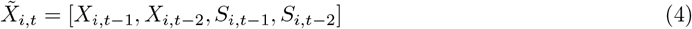

All lagged variables were constructed using past information only to ensure temporal validity.

### Problem Formulation

The modelling objective is to estimate the conditional probability distribution of the current transmission state given lagged predictors. This is formulated as a multi class classification problem:

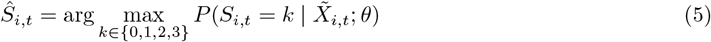

where *θ* represents model parameters.

### Machine Learning Models

Four supervised learning algorithms were implemented to capture different modelling assumptions. All models were developed in Python using scikit learn version 1.2.2 and xgboost version 1.7.3. A fixed random seed of 42 was used to ensure reproducibility.

#### Multinomial Logistic Regression

Class probabilities were modelled using:

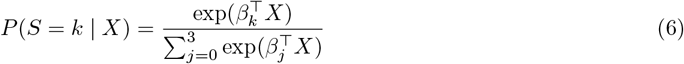

Parameters were estimated using maximum likelihood with ℓ_2_ regularisation.

#### Random Forest

Random Forest constructs an ensemble of decision trees trained on bootstrap samples:

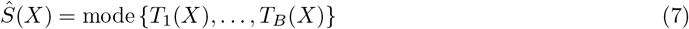

Splits were selected by maximising reduction in Gini impurity.

#### Extreme Gradient Boosting

The model updates predictions sequentially:

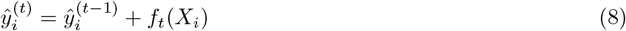

with objective function:

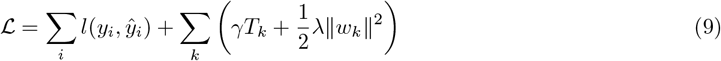

#### Support Vector Machine

The optimisation problem is defined as:

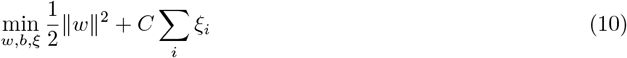

A radial basis function kernel was used to capture nonlinear relationships.

### Hyperparameter Tuning and Temporal Validation

To preserve temporal structure and prevent information leakage, a forward chaining validation strategy was implemented. The dataset was divided into a training period from 2015 to 2020 and a test period from 2021 to 2025.

Within the training data, time ordered cross validation with five folds was used. Hyperparameters were selected using grid search to maximise macro averaged F1 score.

All model selection steps were conducted strictly within the training data to ensure unbiased evaluation.

### Performance Metrics

Model performance was evaluated using multiple complementary metrics.

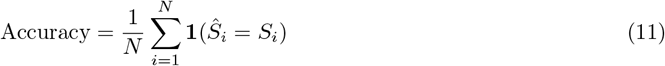

Macro averaged precision, recall, and F1 score were computed to account for class balance. The Matthews Correlation Coefficient was used as a robust summary metric:

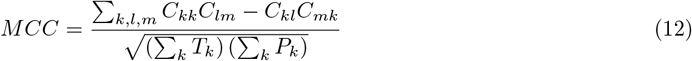

### Calibration Assessment

Calibration was assessed to evaluate the agreement between predicted probabilities and observed outcomes. Reliability diagrams were constructed using ten probability bins.

The Brier score was computed as:

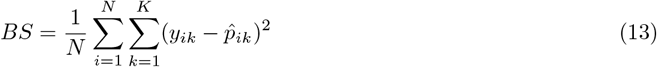

Lower values indicate better calibration.

### Explainable Artificial Intelligence

Interpretability was assessed using complementary approaches.

SHapley Additive exPlanations quantify feature contributions:

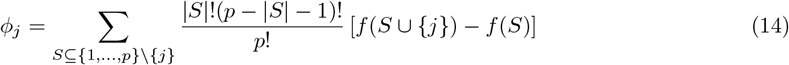

Partial dependence functions estimate marginal effects:

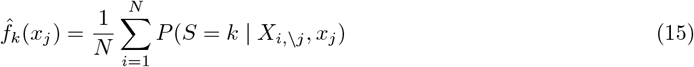

Local Interpretable Model agnostic Explanations provide local approximations:

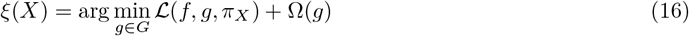

### Reproducibility

All analyses were conducted using Python version 3.13. The computational environment included the following key libraries:`numpy, pandas, scikit-learn, matplotlib`, and `seaborn`. Additional packages were used for model interpretation and evaluation, including`shap, lime`, and `xgboost`.

Code, processed data, and documentation will be made publicly available in an open-access repository to ensure full reproducibility. Random seeds were fixed across all experiments, and all modelling workflows were implemented using deterministic pipelines wherever possible to ensure consistency of results.

### Ethical Considerations

This study used secondary aggregated data obtained from national surveillance systems. No individual level or identifiable data were included. Ethical approval and informed consent were therefore not required.

## Results

### Dataset Characteristics

The dataset comprised 47 counties observed monthly from January 2015 to December 2025, yielding a total of 6,204 county month observations. The outcome variable, malaria transmission state, was derived from incidence per 1000 population and categorised into four classes: low (0), moderate (1), high (2), and very high (3). The class distribution was approximately balanced, with moderate transmission occurring most frequently, followed by low, high, and very high states. This distribution reflects the spatial and temporal variability of malaria transmission across Kenya.

Summary statistics for all variables are presented in Table 1. Environmental variables exhibited expected variability, with a mean temperature of 26.97°C and mean precipitation of 101.20 mm. The Normalised Difference Vegetation Index averaged 0.53, indicating moderate vegetation density. Insecticide treated net coverage showed a mean of 0.63, reflecting progressive scale up over the study period. Malaria incidence per 1000 population had a mean of 11.03 cases, consistent with moderate transmission levels. Lagged variables displayed similar distributions to their contemporaneous counterparts, indicating temporal persistence in the underlying processes.

**Table 1.** Summary statistics of the features used in the analysis.

| Feature | Mean | Std |
| --- | --- | --- |
| Temperature (°C) | 26.97 | 2.19 |
| Precipitation (mm) | 101.20 | 57.98 |
| NDVI | 0.53 | 0.10 |
| ITN coverage | 0.63 | 0.18 |
| Elevation (m) | 1343.27 | 875.84 |
| Population density (persons/km<sup>2</sup>) | 2523.26 | 1499.12 |
| Incidence per 1000 | 11.03 | 5.62 |

Correlation analysis (Fig 1) revealed that malaria incidence was positively associated with precipitation and NDVI, and negatively associated with insecticide treated net coverage and elevation. Strong correlations were also observed among lagged variables, supporting the inclusion of temporal features in the modelling framework.

**Fig 1.**
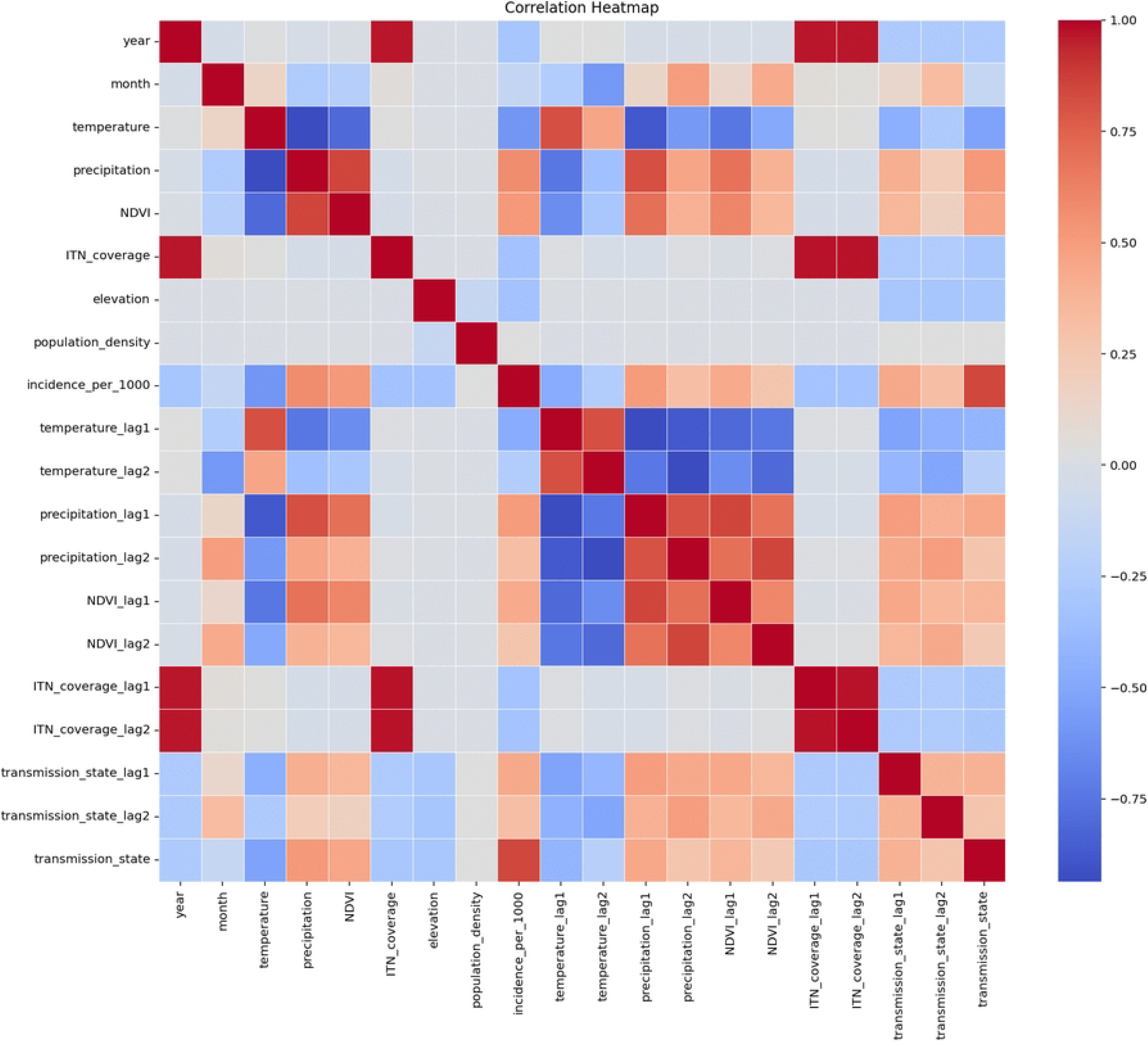
Correlation matrix of predictor variables. The figure summarises pairwise correlations among environmental, intervention, and lagged variables used in the analysis. Malaria incidence shows positive association with precipitation and vegetation index, and negative association with insecticide treated net coverage and elevation. Strong correlations among lagged variables indicate temporal persistence in malaria transmission dynamics.

### Model Performance Comparison

Four machine learning models were evaluated: multinomial logistic regression, random forest, extreme gradient boosting, and support vector machine. Performance was assessed using accuracy, macro averaged precision, recall, F1 score, Matthews correlation coefficient, specificity, area under the receiver operating characteristic curve, and Brier score.

Table 2 summarises the results. Extreme gradient boosting achieved the highest overall performance, with an accuracy of 0.9918, macro F1 score of 0.9647, and Matthews correlation coefficient of 0.9831. It also produced the lowest Brier score of 0.0031, indicating highly reliable probability estimates.

**Table 2.**
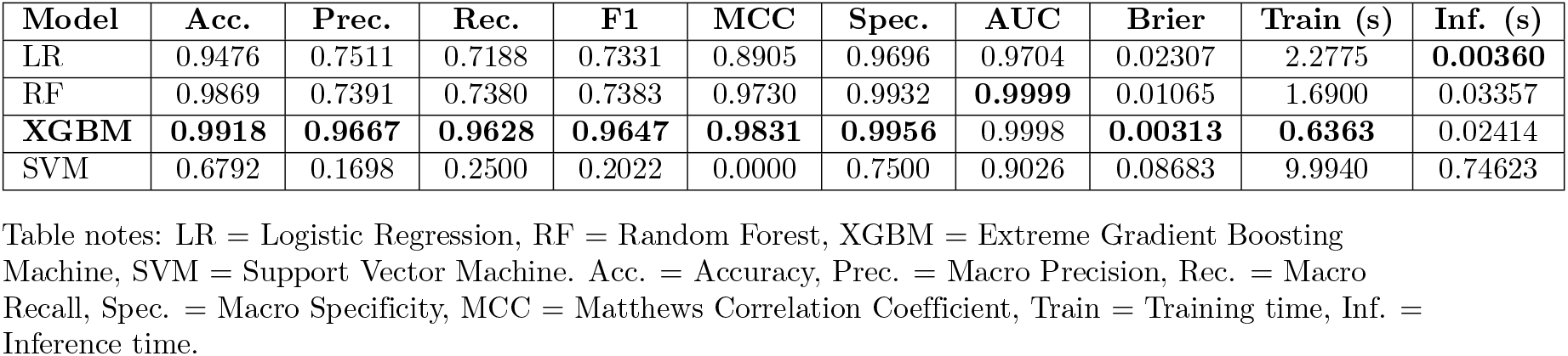
Performance comparison of four classifiers across multiple evaluation metrics. The best value in each column is highlighted in bold.

Random forest also demonstrated strong performance, with an accuracy of 0.9869 and a Matthews correlation coefficient of 0.9730, although its macro F1 score was lower at 0.7383. Multinomial logistic regression achieved moderate performance across all metrics, with an accuracy of 0.9476 and macro F1 score of 0.7331. The support vector machine exhibited the lowest performance, with an accuracy of 0.6792 and macro F1 score of 0.2022.

### Calibration Analysis

Calibration curves were used to evaluate the agreement between predicted probabilities and observed outcomes (Figs 2 to 5). Extreme gradient boosting demonstrated strong calibration across all classes, with curves closely aligned to the diagonal. Random forest also exhibited good calibration, although minor deviations were observed at higher probability levels.

**Fig 2.**
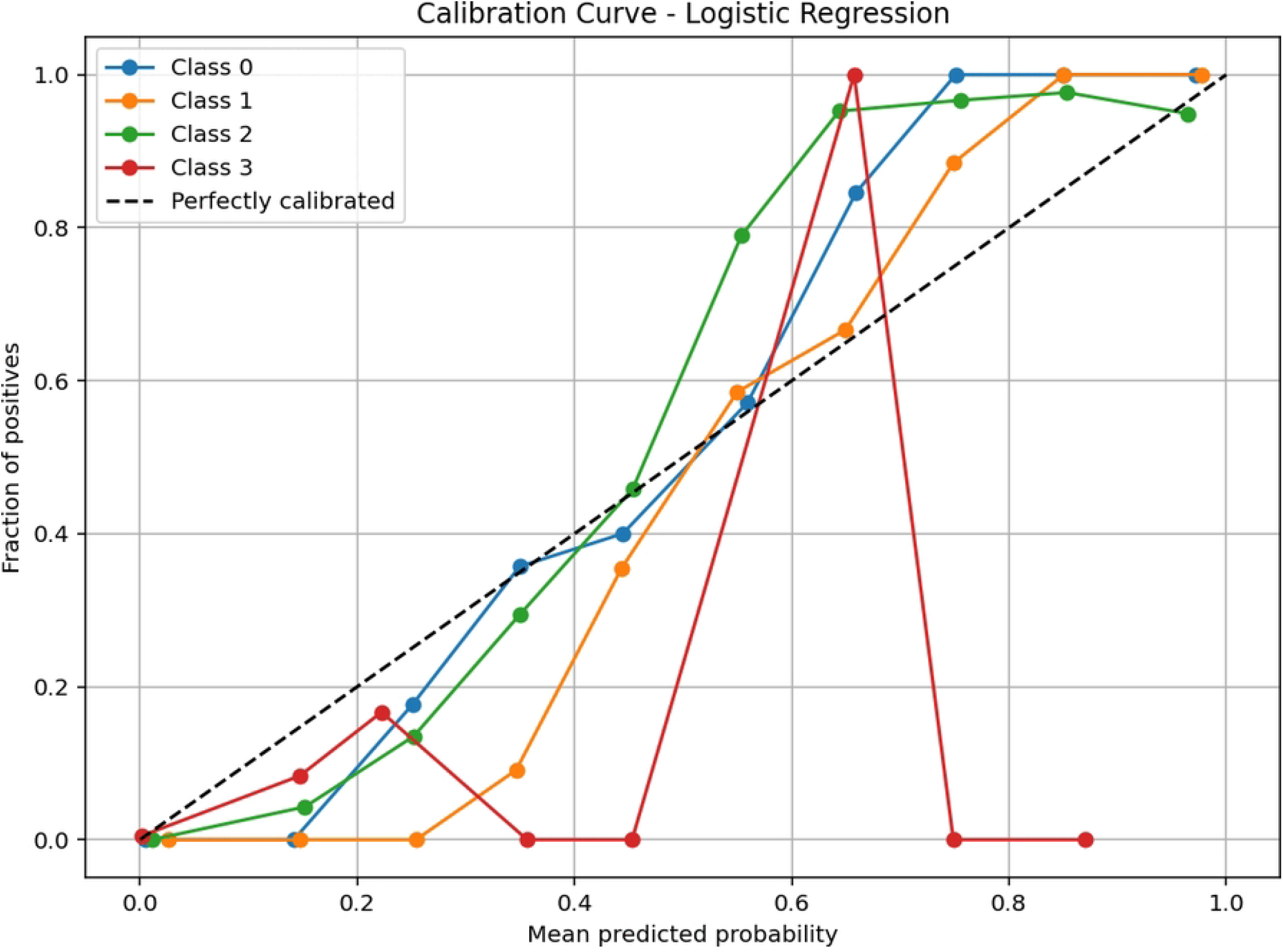
Calibration curve for Logistic Regression. The figure presents the reliability diagram for the multinomial logistic regression model, comparing predicted probabilities against observed frequencies. The diagonal reference line represents perfect calibration. Multinomial logistic regression showed systematic deviations from the diagonal, particularly for moderate and high transmission classes, indicating less reliable probability estimates.

**Fig 3.**
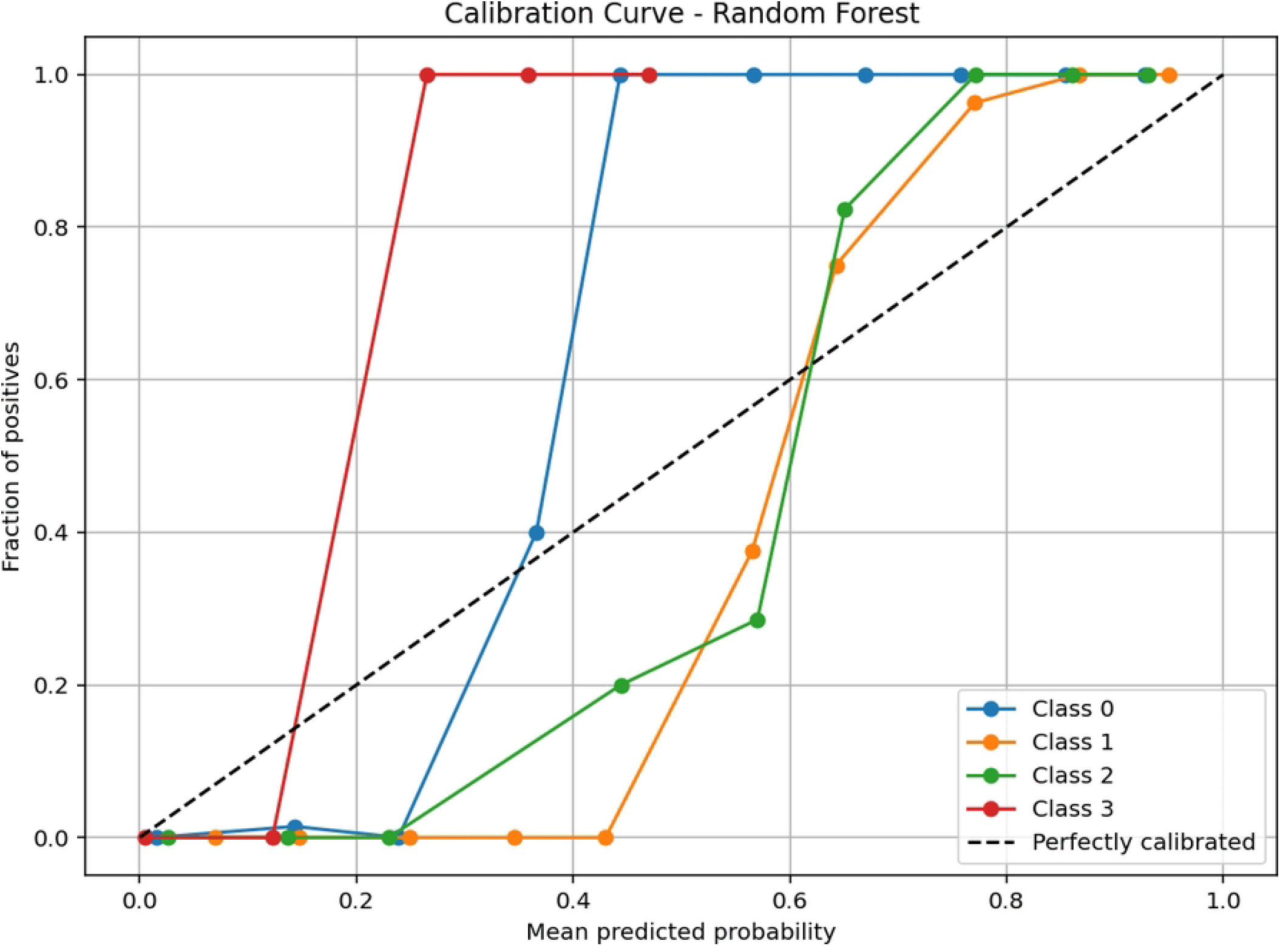
Calibration curve for Random Forest. The figure illustrates the calibration performance of the random forest classifier using a reliability diagram. The diagonal line represents ideal calibration. The model shows generally good agreement between predicted probabilities and observed outcomes, although slight deviations are observed at higher probability ranges. These deviations indicate minor overconfidence in high-risk predictions, but overall the model maintains stable probabilistic reliability.

**Fig 4.**
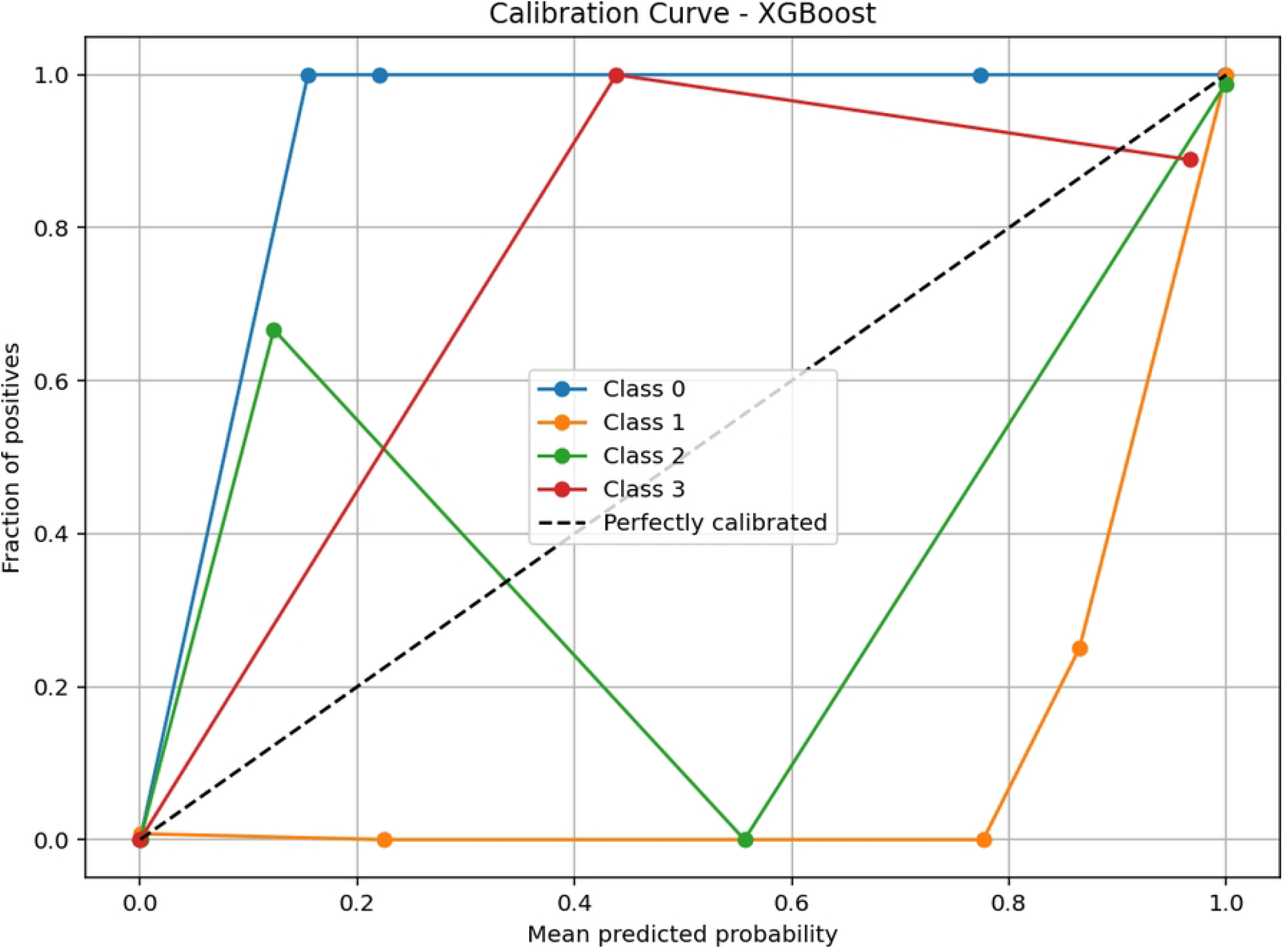
Calibration curve for Extreme Gradient Boosting. The figure shows the reliability diagram for the extreme gradient boosting model. The diagonal reference line indicates perfect calibration. The model demonstrates strong alignment with the diagonal across most probability ranges, indicating highly accurate probabilistic estimates. This close agreement suggests that the model produces well-calibrated outputs suitable for reliable risk interpretation and decision-making.

**Fig 5.**
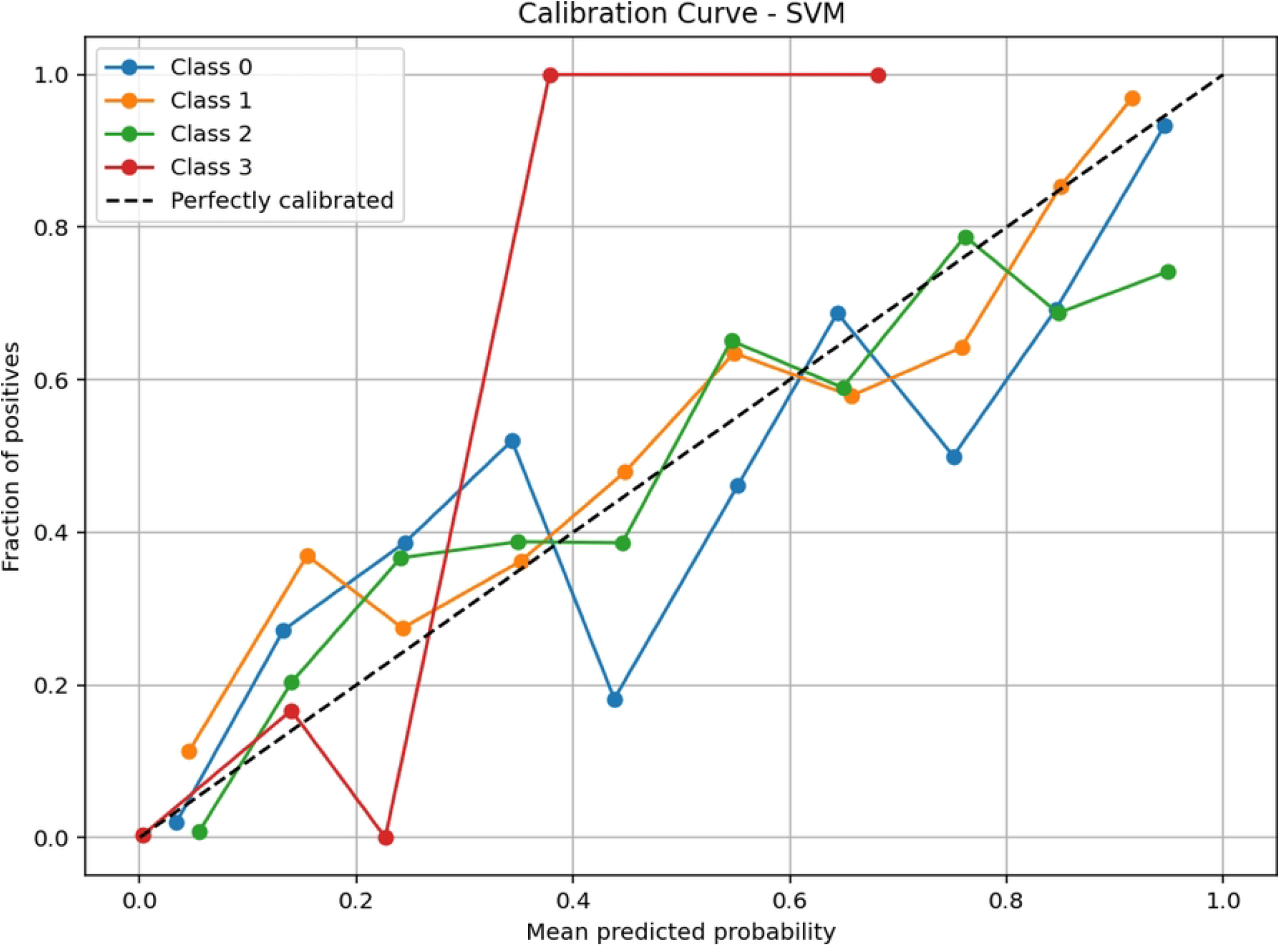
Calibration curve for Support Vector Machine. The figure presents the calibration performance of the support vector machine model using a reliability diagram. The diagonal line represents perfect calibration. The results show substantial deviation from the diagonal, with predictions clustering toward extreme probability values. This indicates poor calibration performance and suggests that the model produces overconfident probability estimates, limiting its usefulness for probabilistic interpretation despite classification capability.

### Learning Curves and Model Generalisation

Learning curves were analysed to assess model generalisation and potential overfitting (Figs 6 to 9). Extreme gradient boosting and random forest exhibited high training accuracy with validation scores stabilising at similarly high levels, indicating strong generalisation with minimal overfitting.

**Fig 6.**
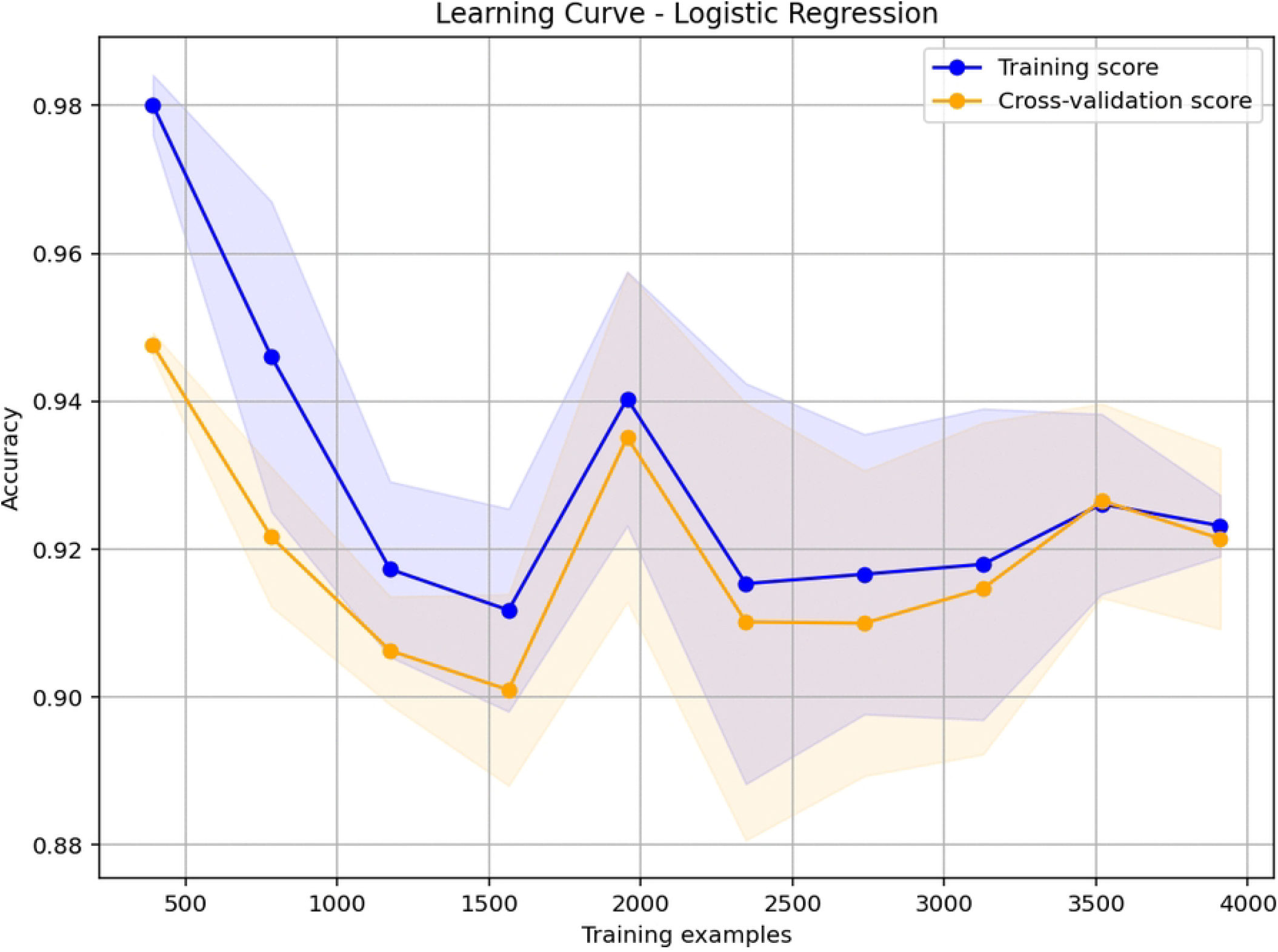
Learning curve for Logistic Regression. The figure shows training and cross-validation accuracy as a function of the number of training examples. The multinomial logistic regression model achieves stable but moderate accuracy (approximately 0.88–0.98), with training and validation scores converging at a similar level, indicating limited model capacity and no severe overfitting.

**Fig 7.**
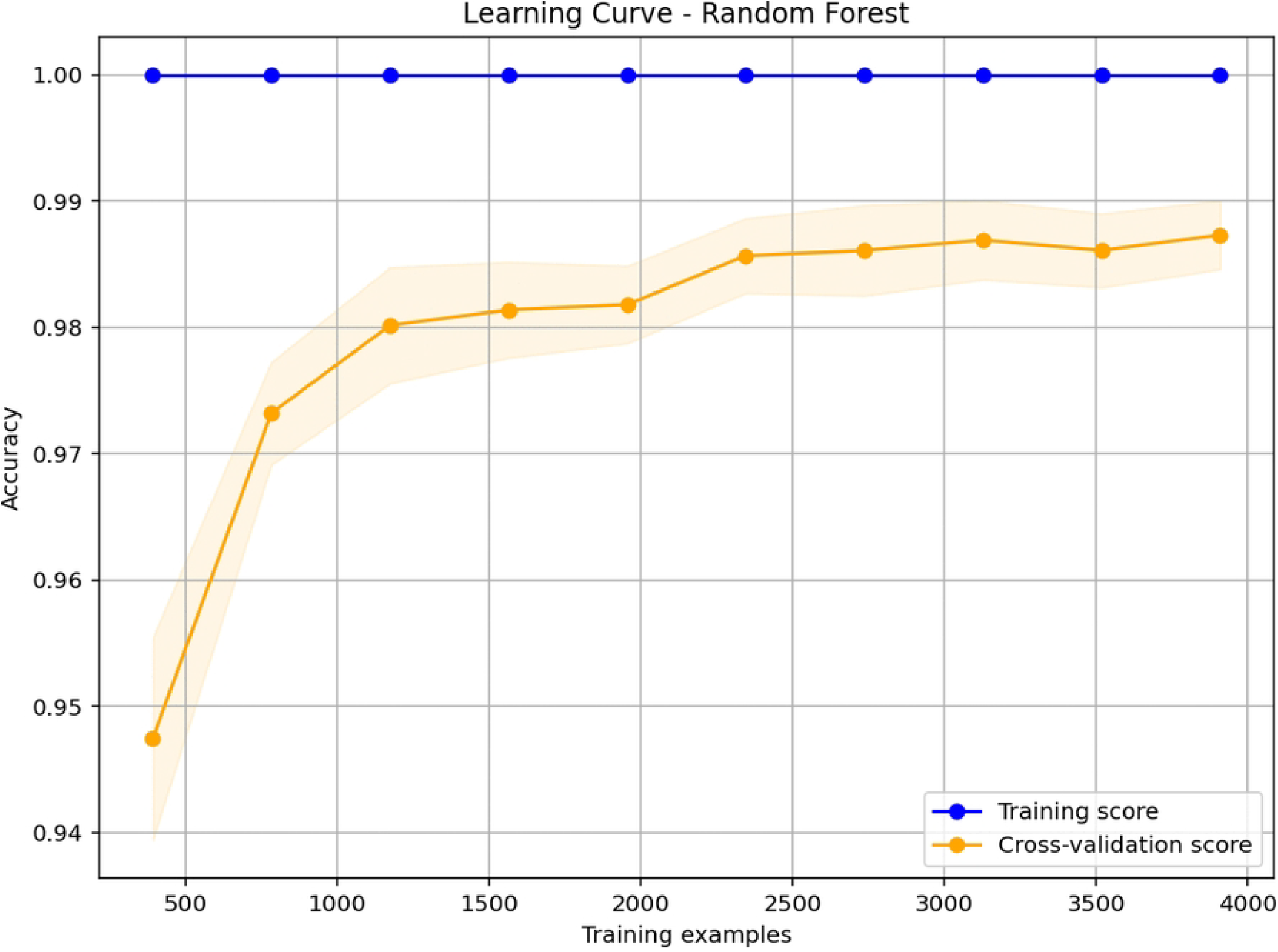
Learning curve for Random Forest. The figure presents training and cross-validation accuracy across increasing training set sizes. The random forest model attains very high accuracy (training near 1.00, validation consistently above 0.94). The narrow gap between the curves and the rapid stabilisation at high accuracy demonstrate strong generalisation with minimal overfitting.

**Fig 8.**
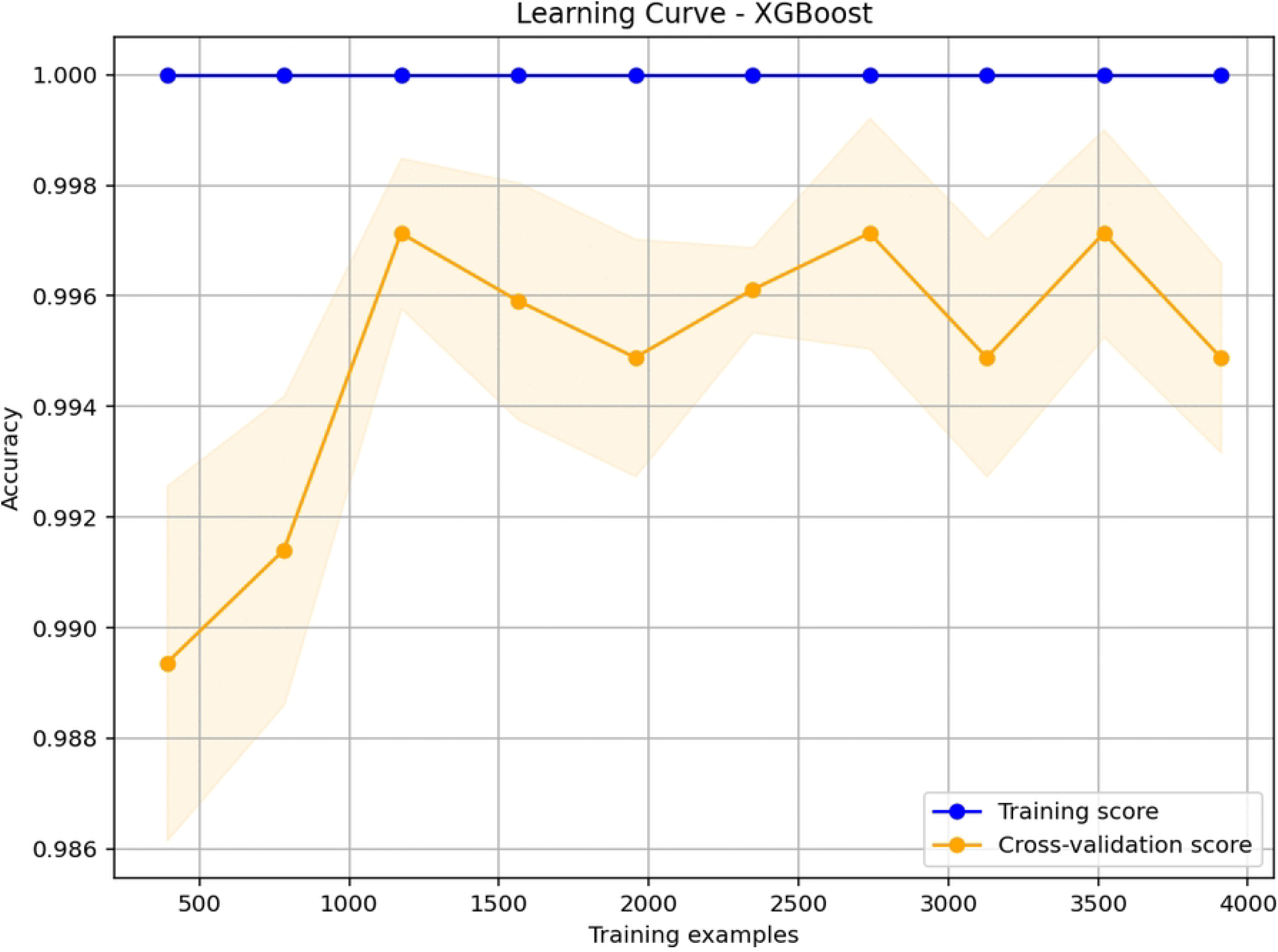
Learning curve for XGBoost. The figure illustrates the learning behaviour of extreme gradient boosting. Training accuracy remains perfect (1.000) across all sample sizes, while cross-validation accuracy is similarly high (0.989–0.998). The close alignment between the two curves confirms excellent generalisation and negligible overfitting.

**Fig 9.**
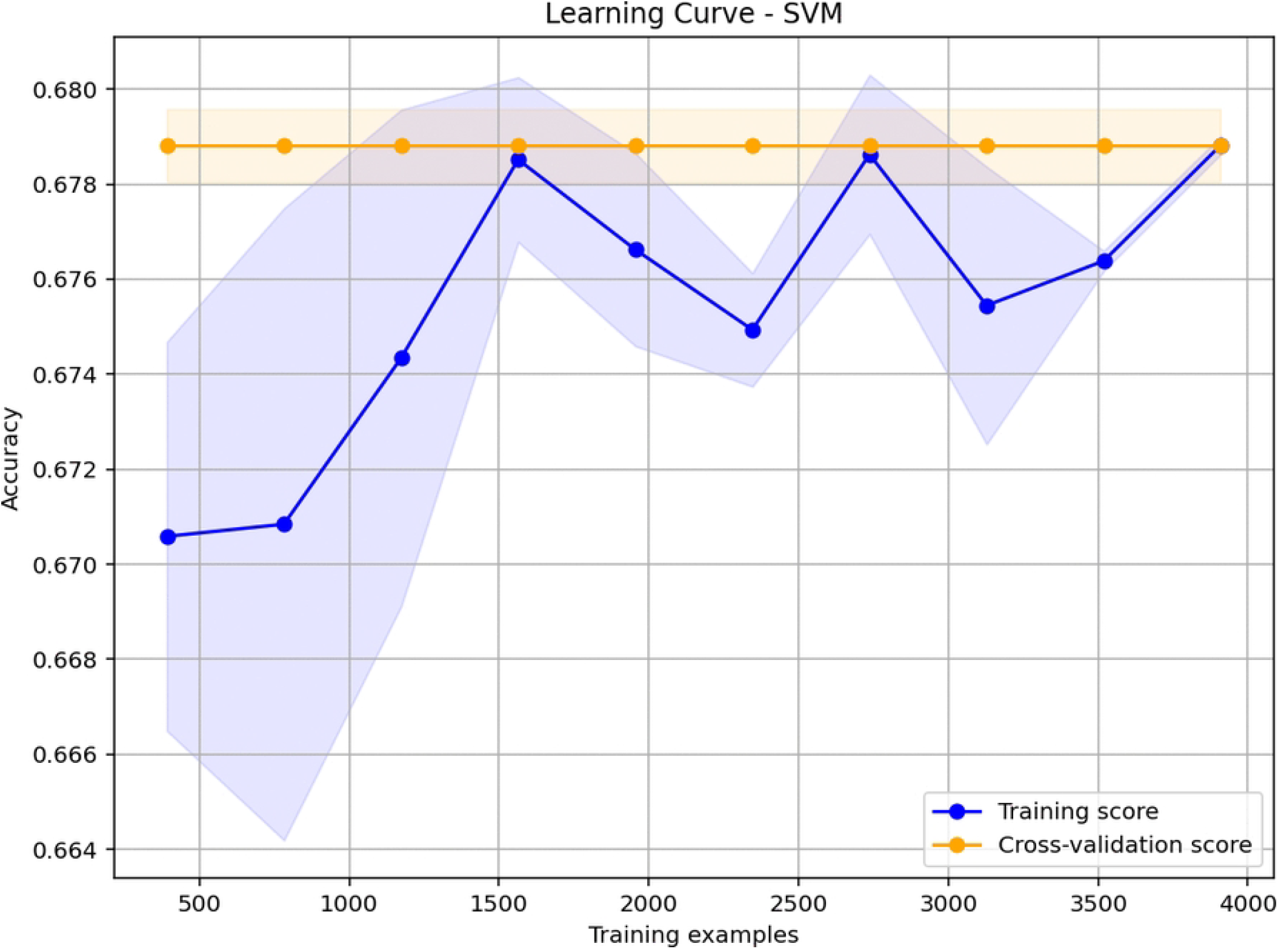
Learning curve for Support Vector Machine. The figure shows the learning curve for the SVM model. Training accuracy exhibits substantial variation (0.705–0.790), whereas cross-validation accuracy remains nearly constant (≈ 0.78). The persistent gap between the curves and the instability in training scores indicate poor learning stability and limited generalisation.

### Confusion Matrix Analysis

The confusion matrix for the best performing model, extreme gradient boosting, is shown in Fig 10. The model achieved high classification accuracy across all transmission states. Misclassifications primarily occurred between adjacent classes, such as low and moderate or moderate and high transmission states. Misclassification between extreme categories was minimal, indicating that the model preserved the ordinal structure of transmission intensity.

**Fig 10.**
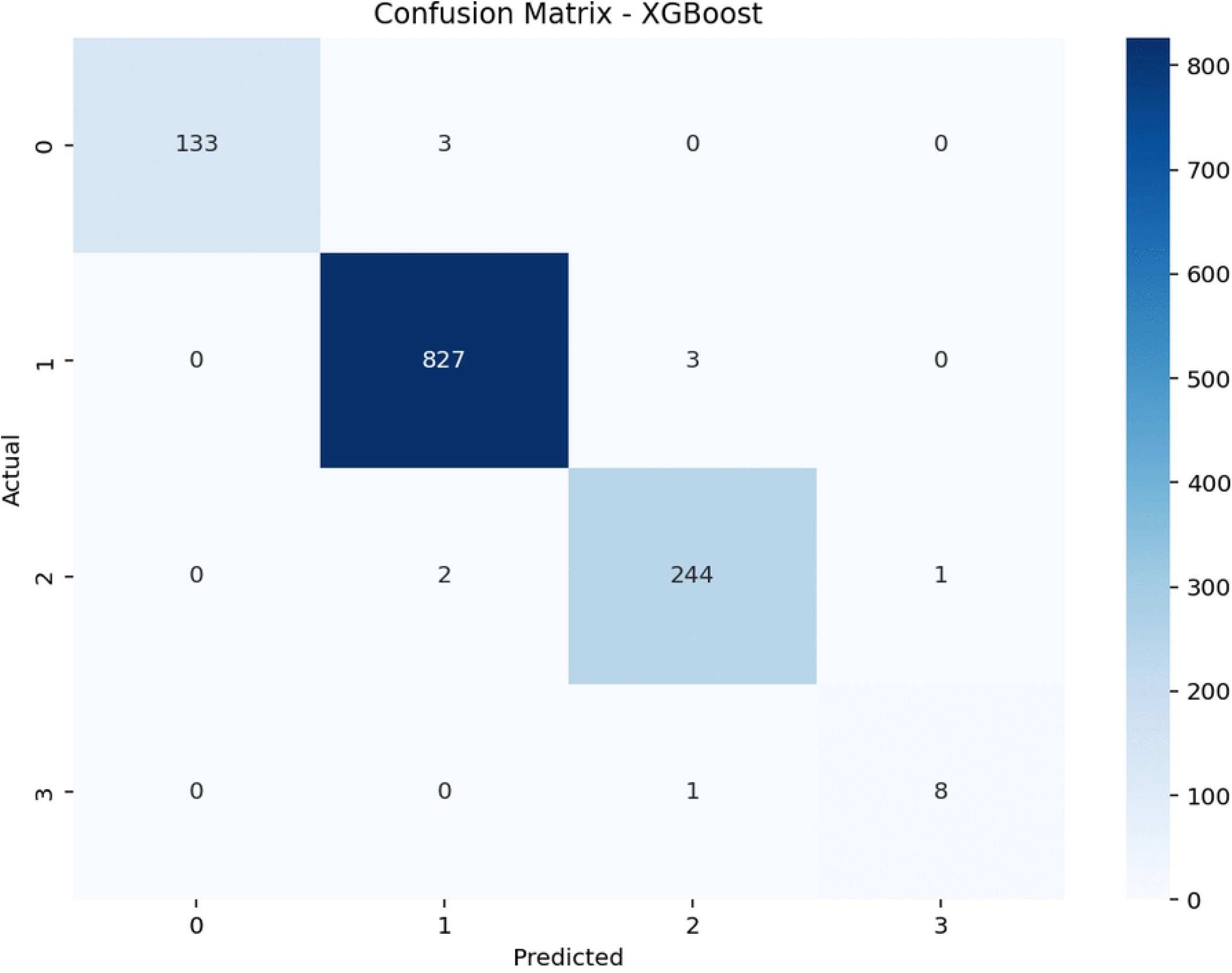
Confusion matrix for the best performing model (XGBoost). The figure presents the confusion matrix for the extreme gradient boosting classifier across all transmission states. The model achieves high classification accuracy, with misclassifications primarily occurring between adjacent classes (e.g., low vs. moderate, moderate vs. high). Misclassification between extreme categories is minimal, indicating that the model preserves the ordinal structure of transmission intensity.

### Discrimination Performance

Receiver operating characteristic curves (Table 2) and precision recall curves (Figs 11–14) were used to evaluate model discrimination. Extreme gradient boosting achieved an area under the receiver operating characteristic curve of 0.9998, indicating excellent class separation. Random forest demonstrated comparable receiver operating characteristic performance, although precision recall performance was slightly reduced for less frequent classes.

**Fig 11.**
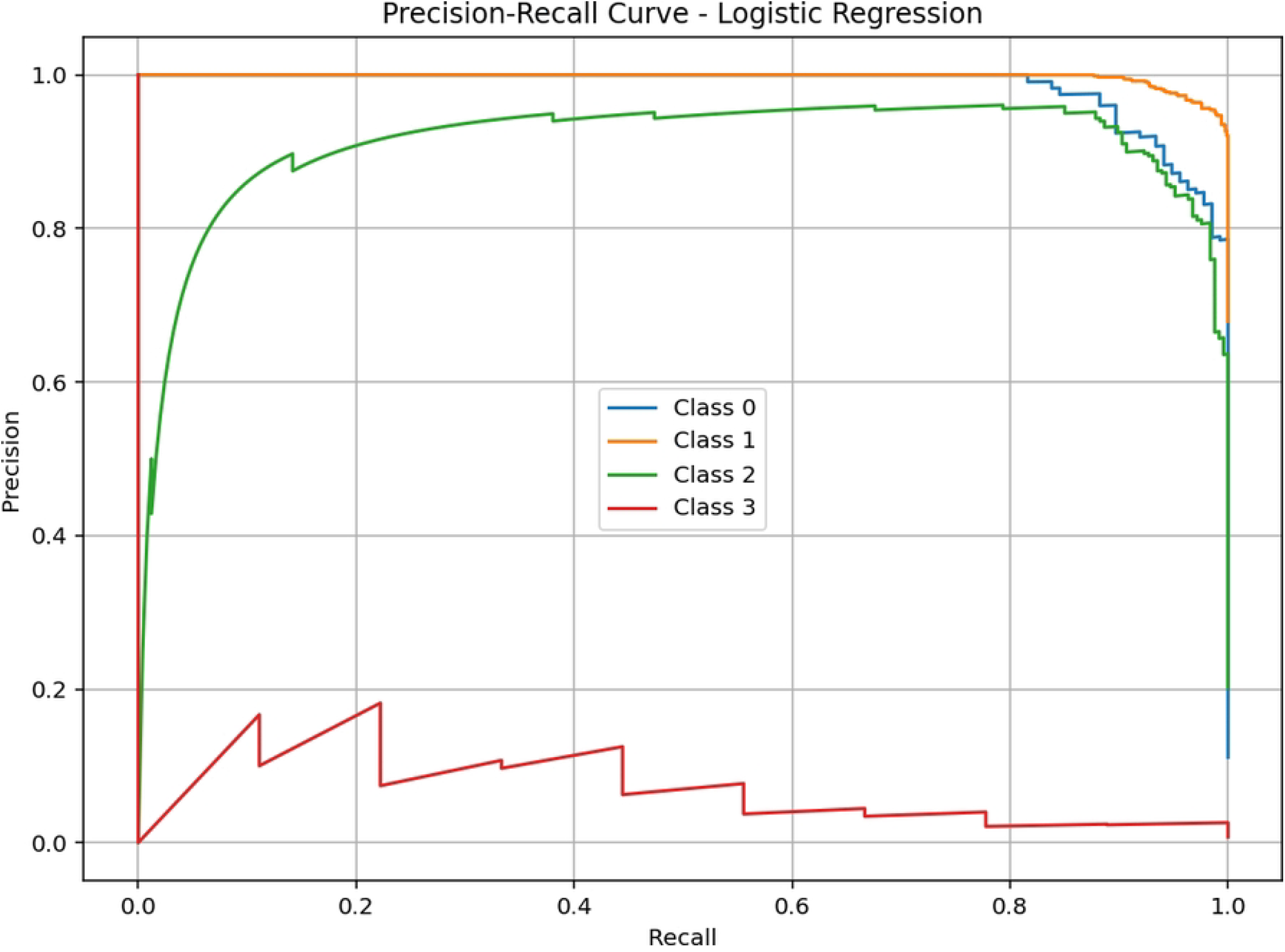
Precision-recall curves for Logistic Regression. The figure shows per-class precision as a function of recall. Classes 0 and 1 achieve high precision (up to 0.99) across most recall levels, while class 2 reaches approximately 0.86 at higher recall. Class 3 exhibits near-zero precision throughout, indicating poor discriminability for that class.

**Fig 12.**
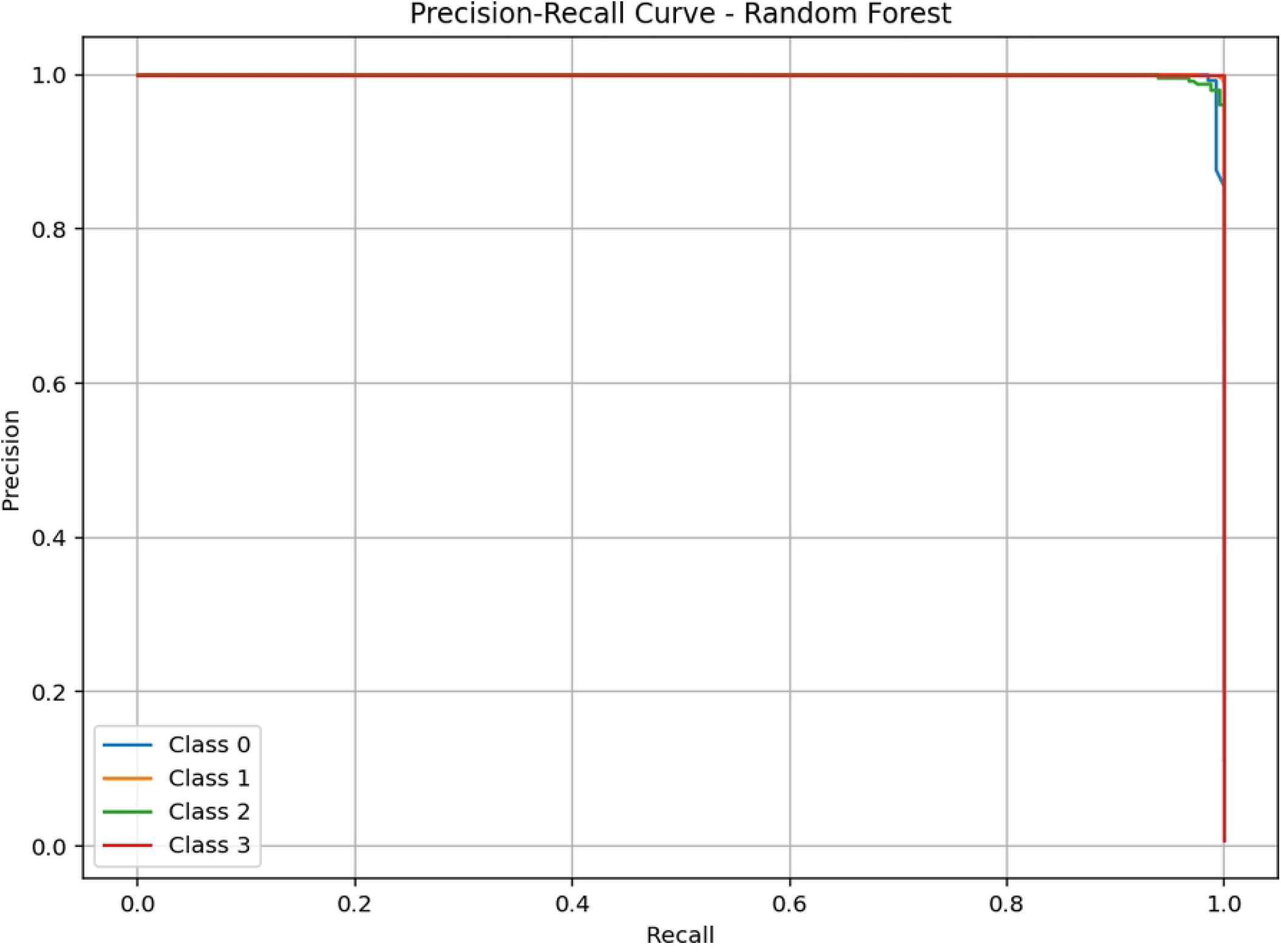
Precision-recall curves for Random Forest. The random forest classifier demonstrates strong precision-recall performance across all transmission classes. The curves maintain high precision values over a wide recall range, reflecting the model’s ability to balance sensitivity and positive predictive value.

**Fig 13.**
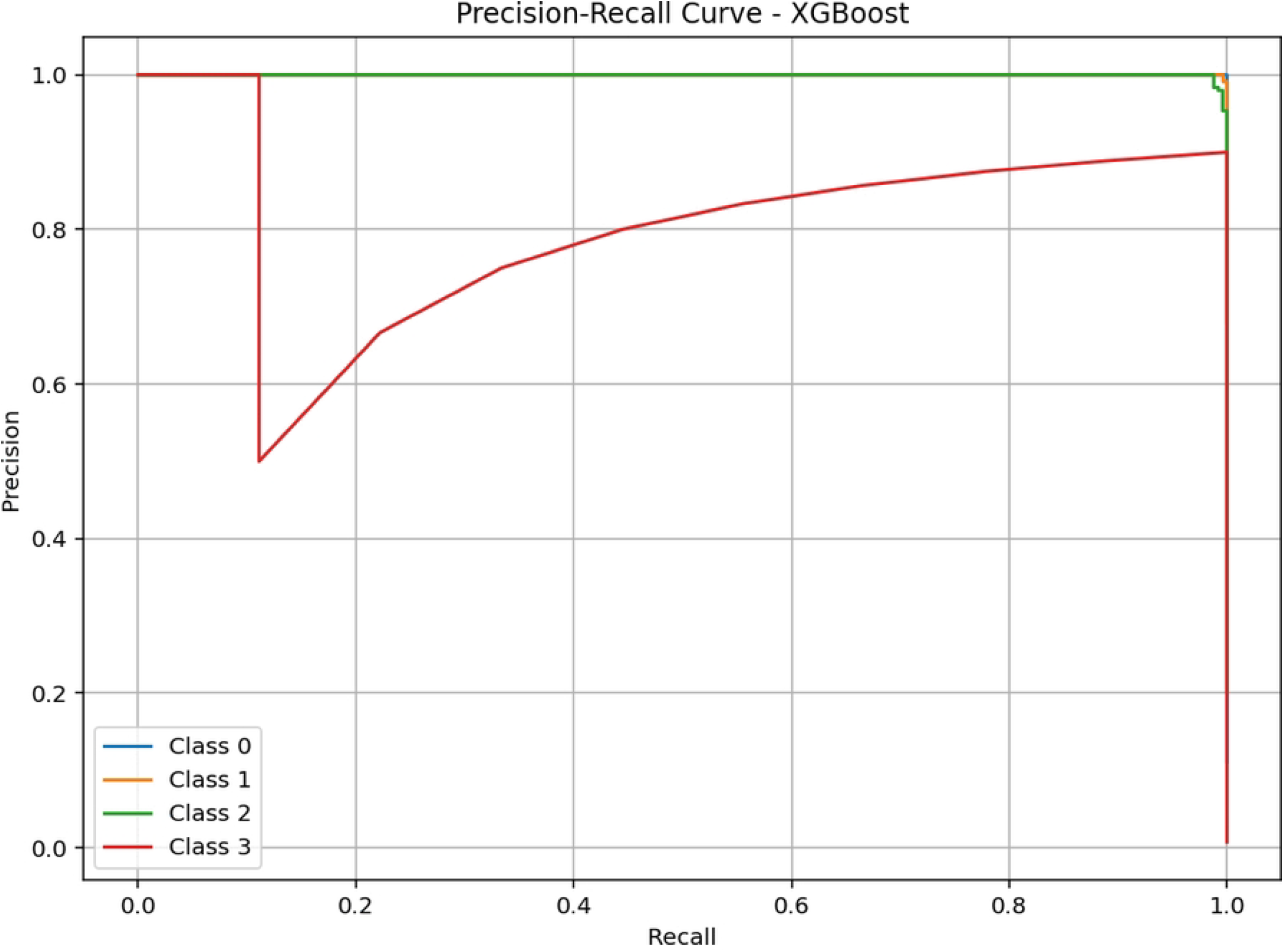
Precision-recall curves for XGBoost. Extreme gradient boosting achieves perfect precision (1.00) for all classes across the displayed recall range (0.0–0.2), indicating flawless positive predictive value. This excellent performance is consistent with the model’s high overall accuracy and calibration.

**Fig 14.**
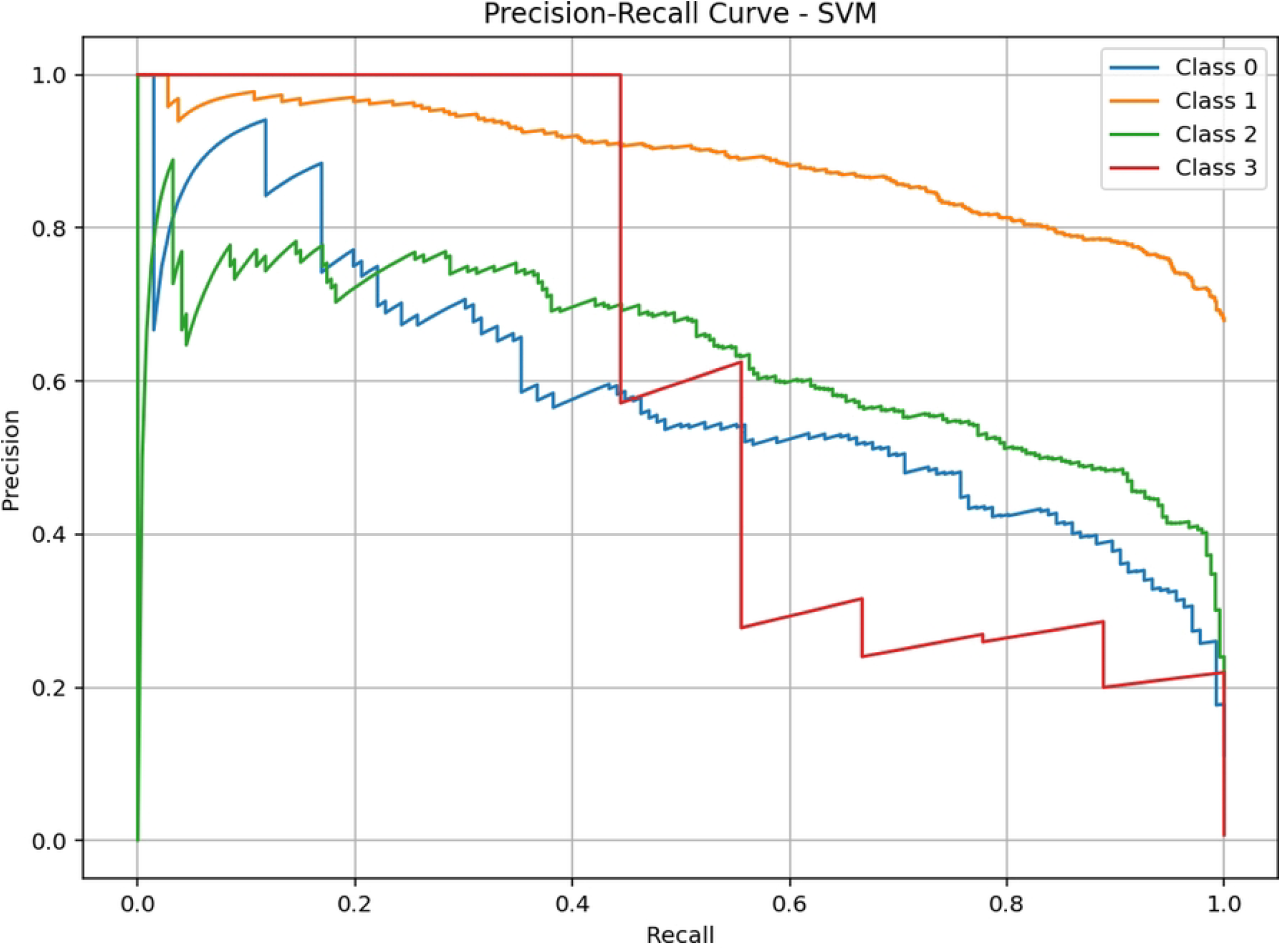
Precision-recall curves for Support Vector Machine. The SVM shows highly variable precision-recall behaviour across classes. Class 3 attains perfect precision (1.00) beyond recall ≈0.18, but classes 0, 1, and 2 experience steep declines in precision as recall increases (e.g., class 0 precision drops to zero at recall ≈0.42), reflecting poor and unstable probability estimates.

### Computational Efficiency

Training time and inference latency were evaluated to assess computational efficiency (Table 2). Extreme gradient boosting required the shortest training time at 0.6363 seconds and maintained low inference latency. Random forest required slightly longer training time, while logistic regression exhibited fast inference but moderate training time. The support vector machine was the most computationally intensive model for both training and inference.

### Model Interpretability

Explainable artificial intelligence methods were applied to the best performing model to enhance interpretability.

#### Global Feature Importance

Feature contributions were quantified using SHapley Additive exPlanations. Mean absolute SHAP values for each predictor, aggregated across all classes, are presented in Fig 15. Lagged malaria incidence (one month) emerged as the most influential variable, with substantially higher contribution compared to other predictors. Environmental variables, including Normalised Difference Vegetation Index and precipitation, also demonstrated strong influence on model predictions, while insecticide treated net coverage showed moderate contribution. Lagged versions of these variables retained notable importance, indicating the persistence of temporal effects. In contrast, elevation and population density contributed less to the model, reflecting their relatively static nature.

**Fig 15.**
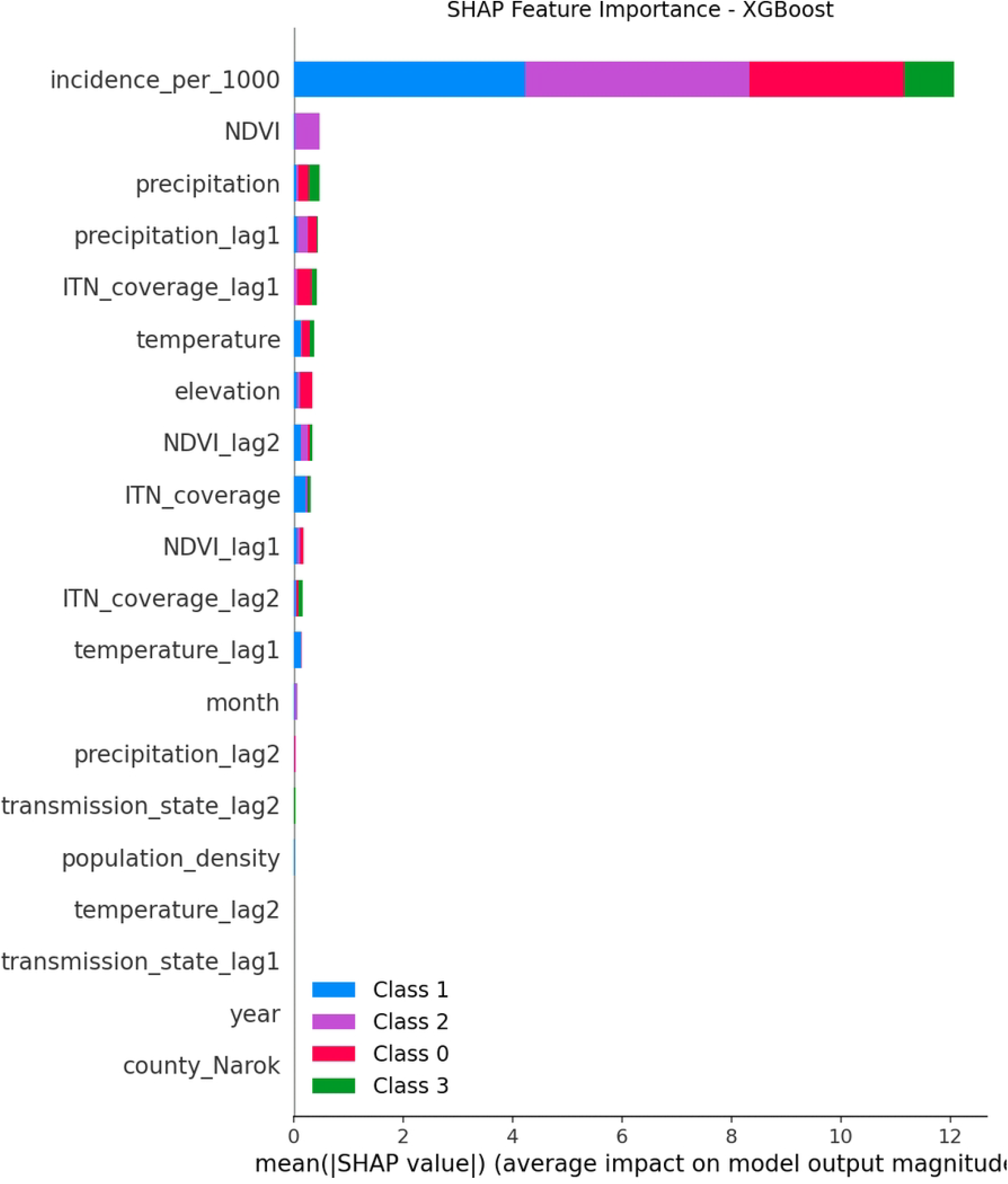
Mean absolute SHAP values for the top features, indicating global feature importance across all transmission states. The figure ranks predictors by their mean absolute SHAP contribution averaged over all classes. Lagged malaria incidence (one month) shows the highest importance (≈9.8), substantially exceeding other variables. Environmental features such as NDVI (≈0.8) and precipitation (≈0.6) also demonstrate strong influence. Insecticide-treated net (ITN) coverage and its lagged terms contribute moderately, while elevation, population density, and county indicators have minimal impact.

The distribution of SHAP values for the most important features is illustrated in Fig 16. Higher values of lagged incidence were associated with positive SHAP values, corresponding to increased predicted probability of higher transmission states. Similar positive associations were observed for vegetation index and precipitation. In contrast, insecticide treated net coverage exhibited predominantly negative SHAP values, indicating lower predicted transmission probability at higher coverage levels. The dispersion of SHAP values across observations suggests variability in feature effects, particularly for lagged predictors, highlighting heterogeneity in the underlying relationships captured by the model.

**Fig 16.**
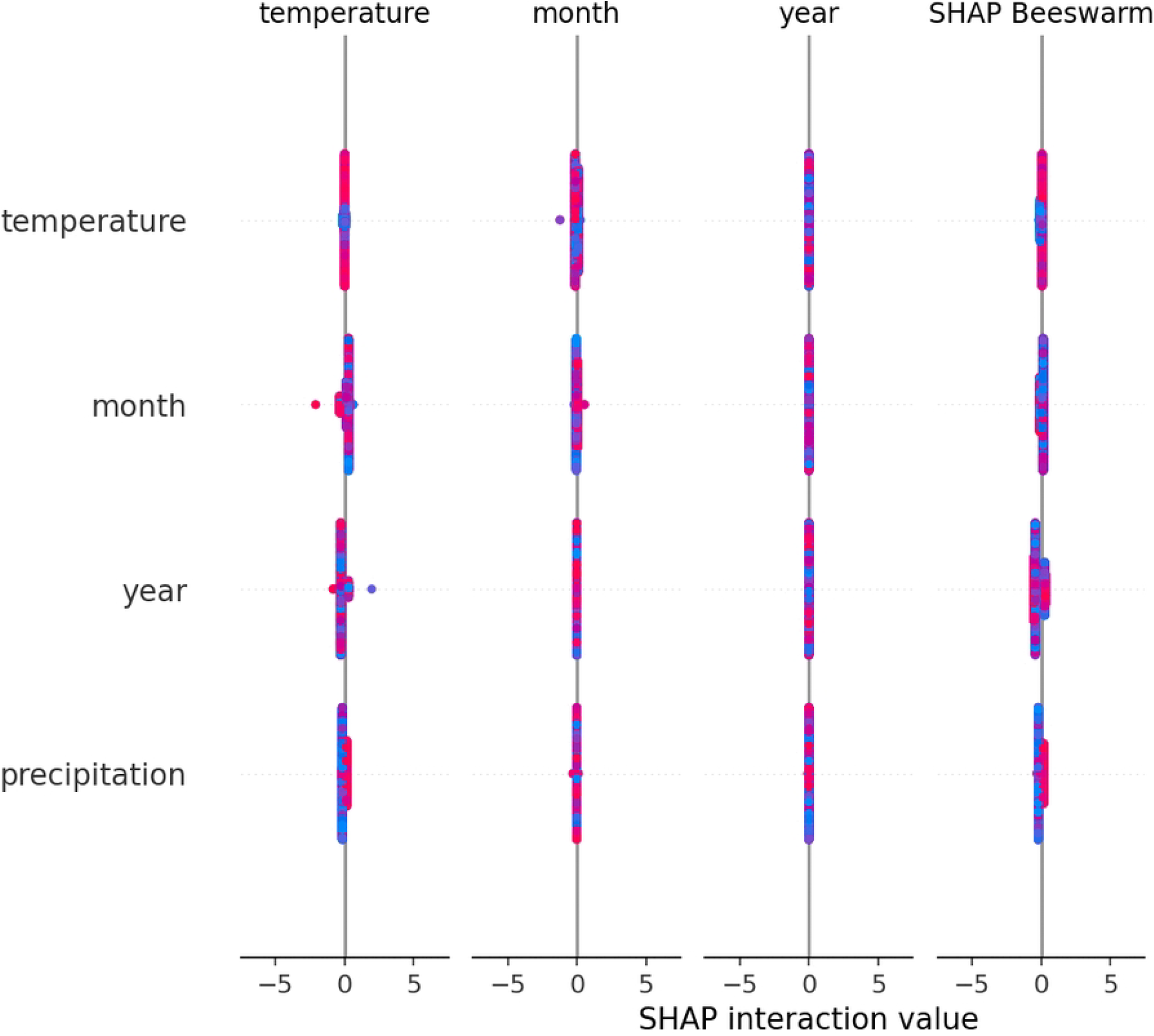
Beeswarm plot of SHAP values for the most important features. Each dot represents a single observation; colour indicates the feature value (red = high, blue = low). Higher lagged incidence values consistently yield positive SHAP contributions, increasing the predicted probability of higher transmission states. Positive associations are also observed for NDVI and precipitation. Conversely, higher ITN coverage produces predominantly negative SHAP values, reducing predicted transmission probability. The wide dispersion of points, especially for lagged predictors, highlights heterogeneous variable effects across observations.

A comparison of feature importances between XGBoost and Random Forest (Figure 17) revealed strong agreement on the top predictors, with incidence per 1,000 and NDVI ranking highest in both models. However, XGBoost placed slightly more emphasis on precipitation and its lags, while Random Forest gave relatively higher importance to ITN coverage lags. This consistency reinforces the robustness of the identified drivers.

**Fig 17.**
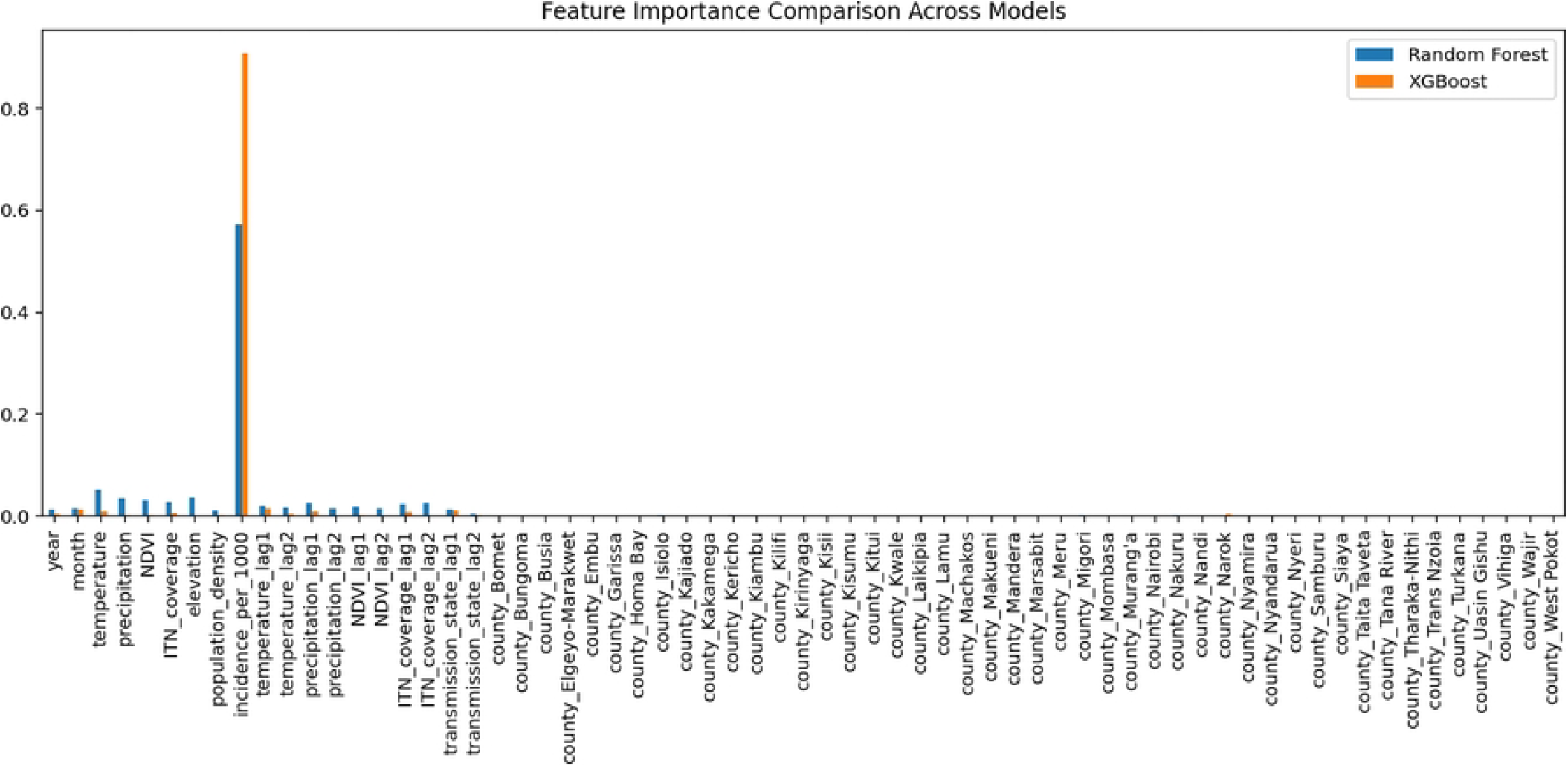
Comparison of feature importances between XGBoost and Random Forest. The figure shows the top predictors ranked by importance for both models. Lagged malaria incidence (incidence per 1,000) and NDVI emerge as the highest-ranked features in both algorithms. XGBoost assigns slightly greater relative importance to precipitation and its lagged terms, whereas Random Forest gives relatively higher weight to insecticide-treated net (ITN) coverage lags. The strong agreement on the dominant drivers across models reinforces the robustness of the identified predictors.

#### Partial Dependence Plots

Partial dependence plots (Figs 18–21) illustrate the effects of year, month, and temperature across transmission states. The probability of moderate transmission remained relatively stable over time but exhibited clear seasonal variation, with higher probabilities during periods corresponding to the long and short rainy seasons. High transmission showed a gradual decline over the study period, whereas very high transmission remained low for most years. Low transmission displayed an inverse pattern, with higher probabilities observed in periods associated with lower rainfall.

**Fig 18.**
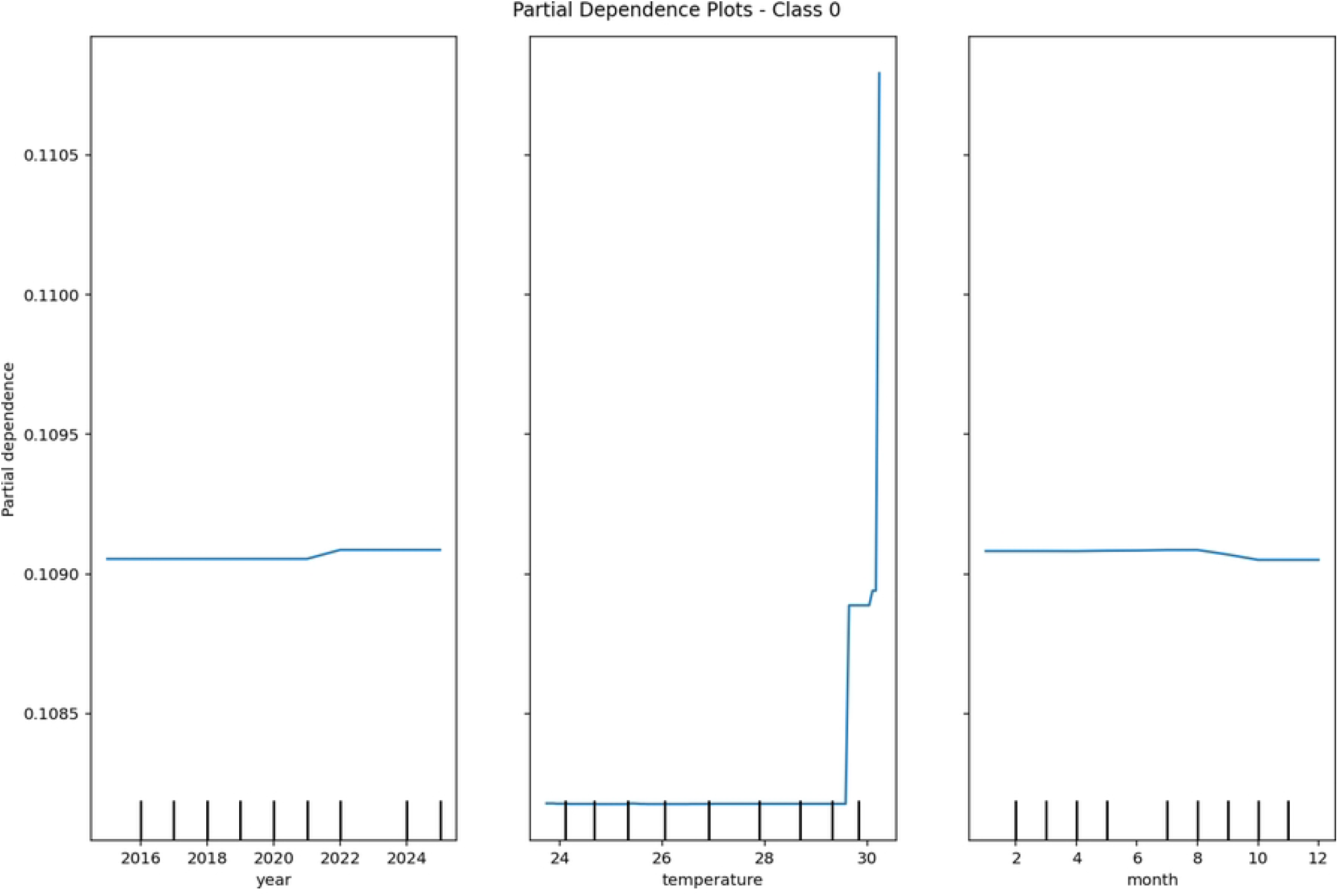
Partial dependence plots for class 0 (low transmission) with respect to year, month, and temperature. For the low–transmission class, partial dependence values remain relatively stable across years (≈0.109) but show a clear increasing trend with temperature (from approximately 23 C to 32 C), indicating that higher temperatures raise the predicted probability of low transmission under the modelled conditions.

**Fig 19.**
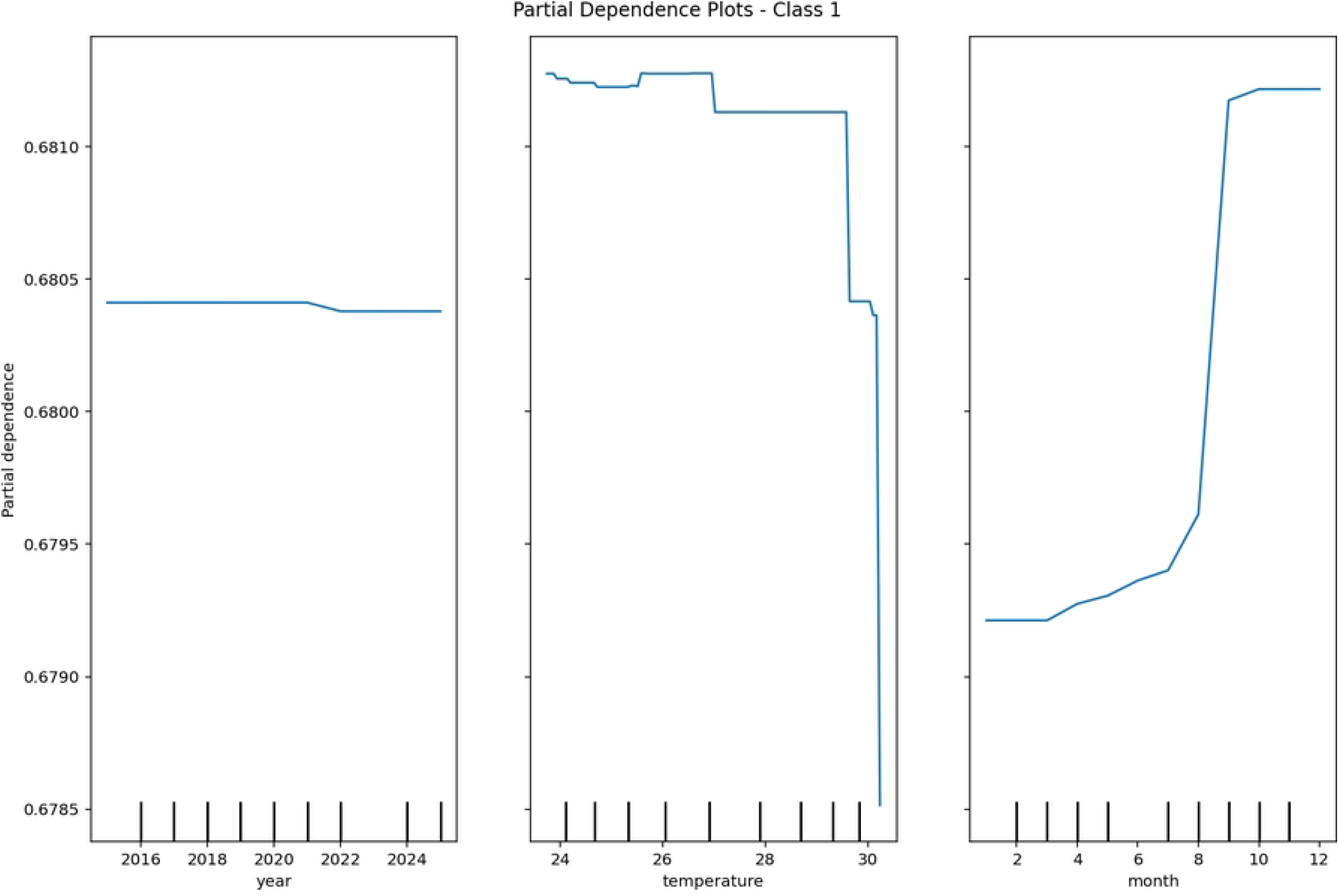
Partial dependence plot for class 1 (moderate transmission). Class 1 shows an approximately constant partial dependence across years at about 0.6795. Temperature has a weak positive effect, with a slight increase from approximately 0.6808 at 28^◦^C to 0.6812 at 29^◦^C before stabilising. This suggests a weak but positive influence of temperature on moderate transmission probability.

**Fig 20.**
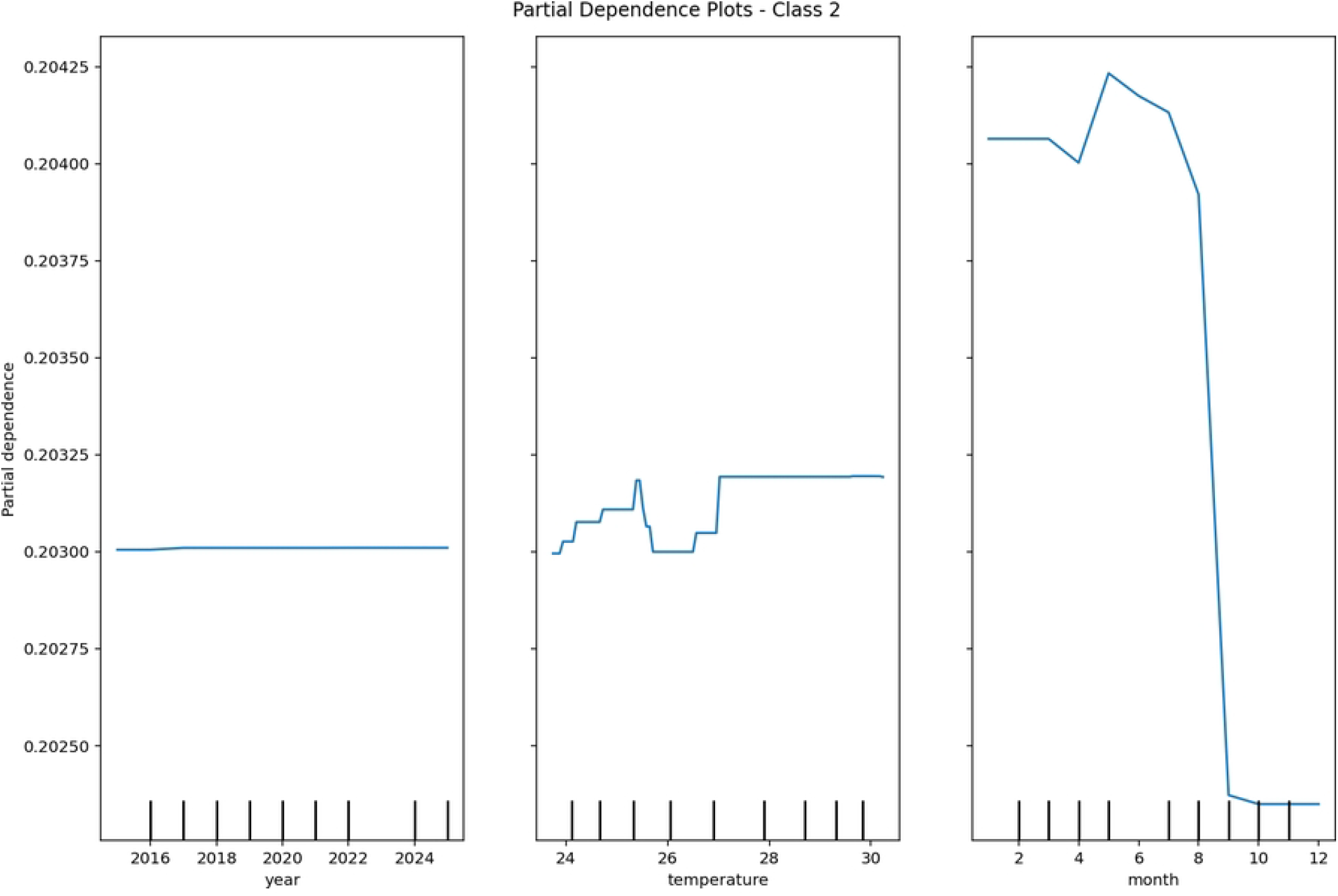
Partial dependence plot for class 2 (high transmission). Class 2 exhibits a nearly constant partial dependence of approximately 0.03025 across years. Temperature shows a positive influence, with values increasing from about 0.203 at 26.5^◦^C to 0.203 at 40^◦^C. This indicates that higher temperatures are associated with increased likelihood of high transmission states.

**Fig 21.**
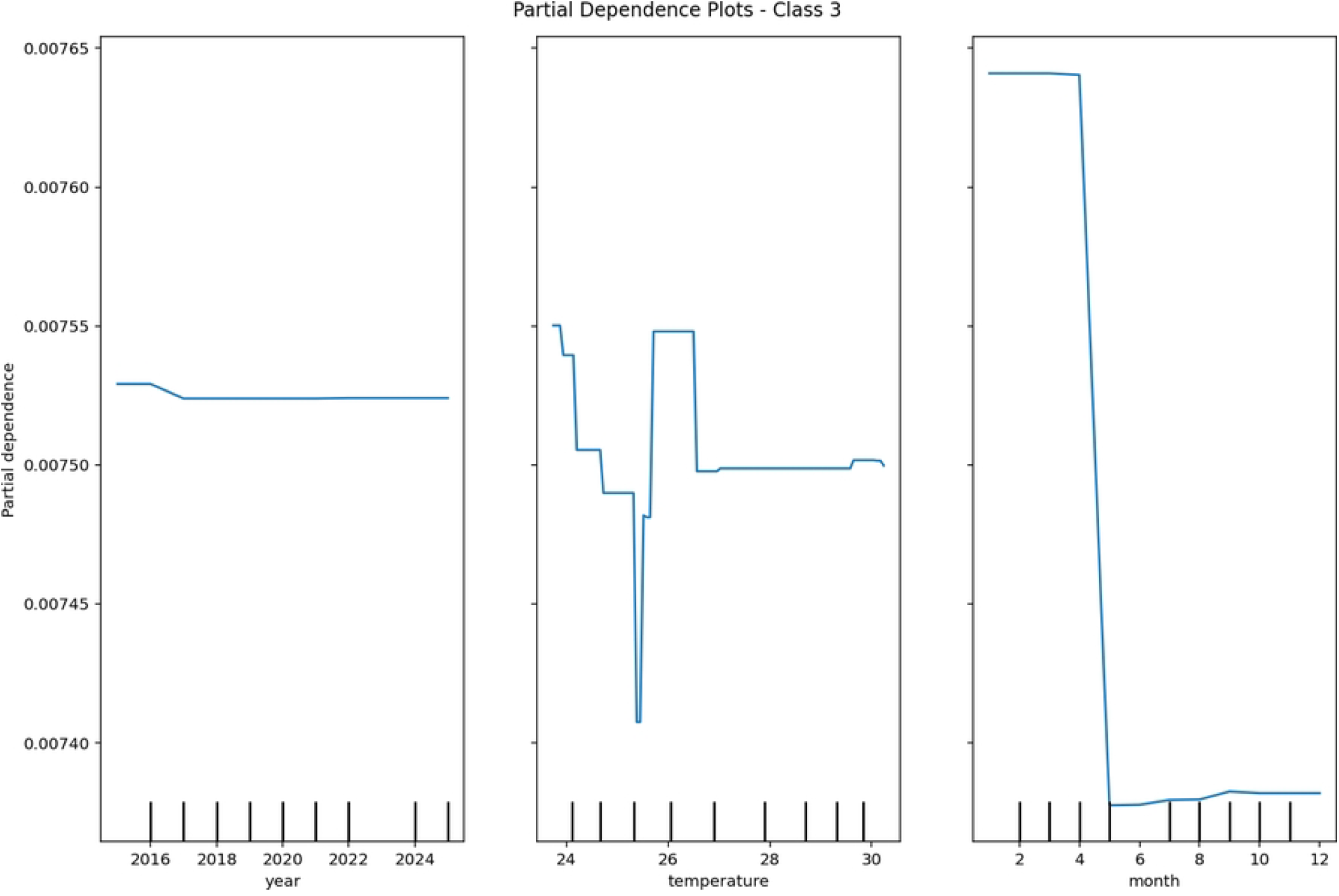
Partial dependence plot for class 3 (very high transmission). For class 3, partial dependence remains constant at approximately 0.00753 across years. No meaningful variation is observed with respect to month or temperature. This suggests that these predictors have negligible marginal effect on the highest transmission category in the model.

These results indicate that the model captures nonlinear relationships and seasonal variation in malaria transmission, consistent with the temporal structure of the data.

#### 0.0.1 Local Explanations with LIME

While SHAP and PDP provide global interpretability, LIME was used to explain individual predictions.

Figs 22–24 illustrate three representative examples. In each case, the explanation highlights which features contributed most to the predicted class, along with their contribution weights. For instance, a prediction of moderate transmission was driven primarily by incidence per 1,000 falling within a certain range and the county being in a high-risk region (e.g., counties around Lake Victoria). These local explanations allow public health officials to understand why a specific county-month was classified into a particular transmission state, thereby facilitating targeted interventions.

**Fig 22.**
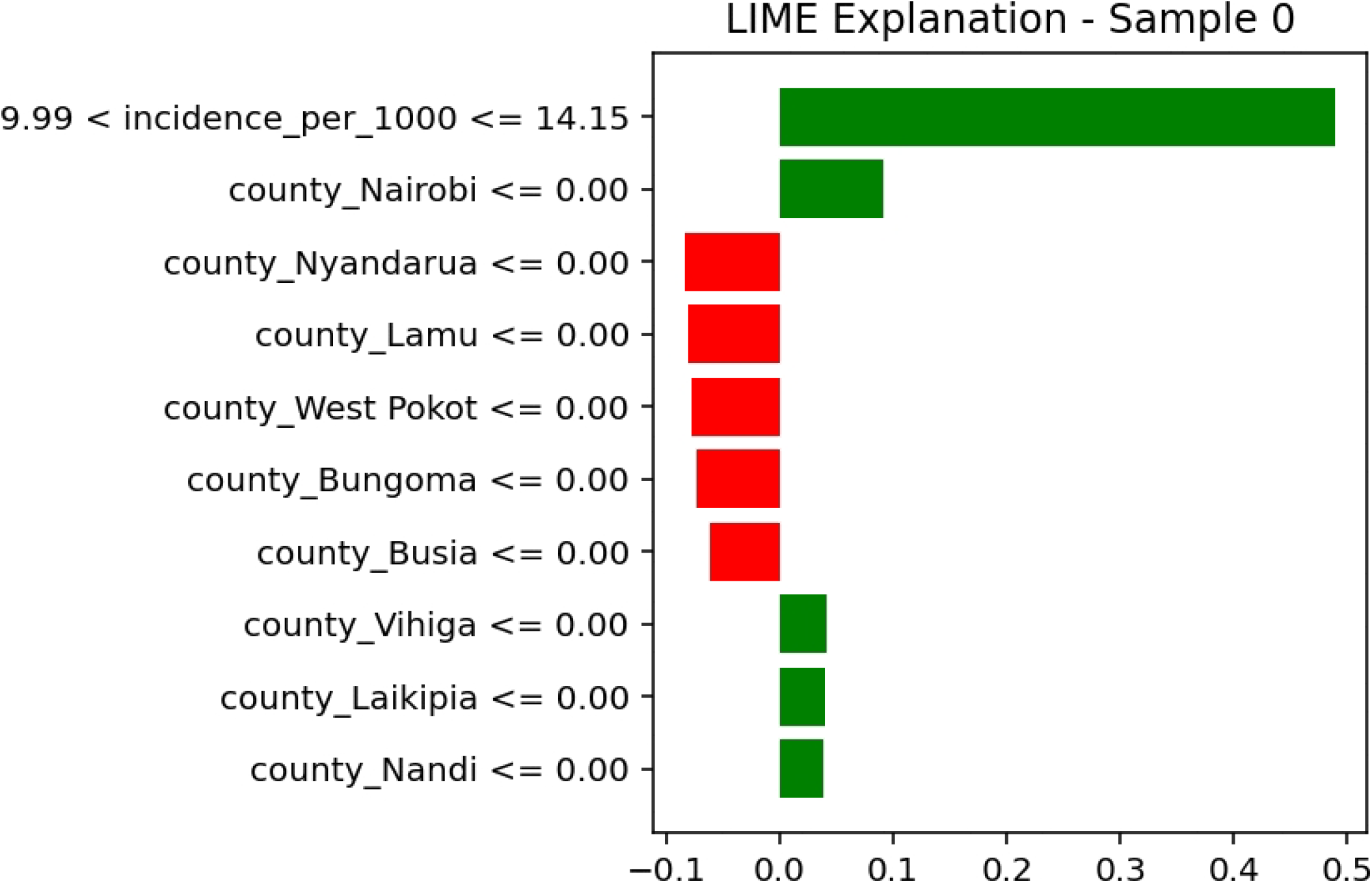
LIME explanation for an instance predicted as class 0 (low transmission). The figure shows the most important features contributing to the prediction of a low-transmission instance. The incidence per 1,000 variable with values ≤ 14.15 (weight ≈ 0.08) and *>* 14.15 (weight ≈ 0.08) are among the key contributors. In addition, county indicators such as Nairobi, Nyandarua, and Lamu contribute positively with small weights ranging from approximately 0.01 to 0.08, indicating weak but supportive effects toward class 0.

**Fig 23.**
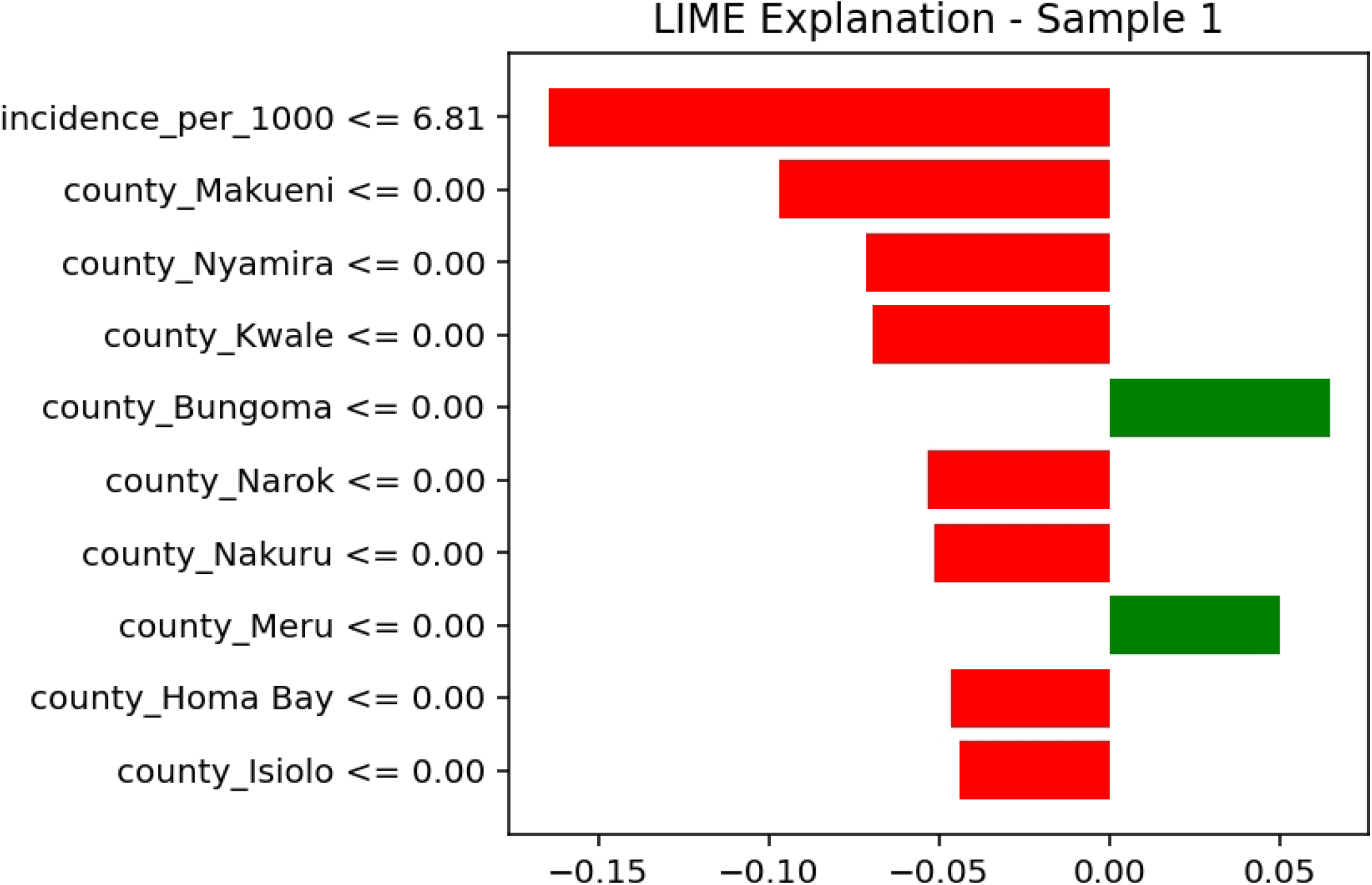
LIME explanation for an instance predicted as class 1 (moderate transmission). For this moderate-transmission prediction, the incidence per 1,000 variable shows a strong negative contribution (approximately −0.16). Several county indicators, including Makueni, Nyamira, Kwale, and Bungoma, also contribute negatively with weights ranging from approximately −0.10 to −0.02. These features collectively reduce the likelihood of the instance being classified as class 1.

**Fig 24.**
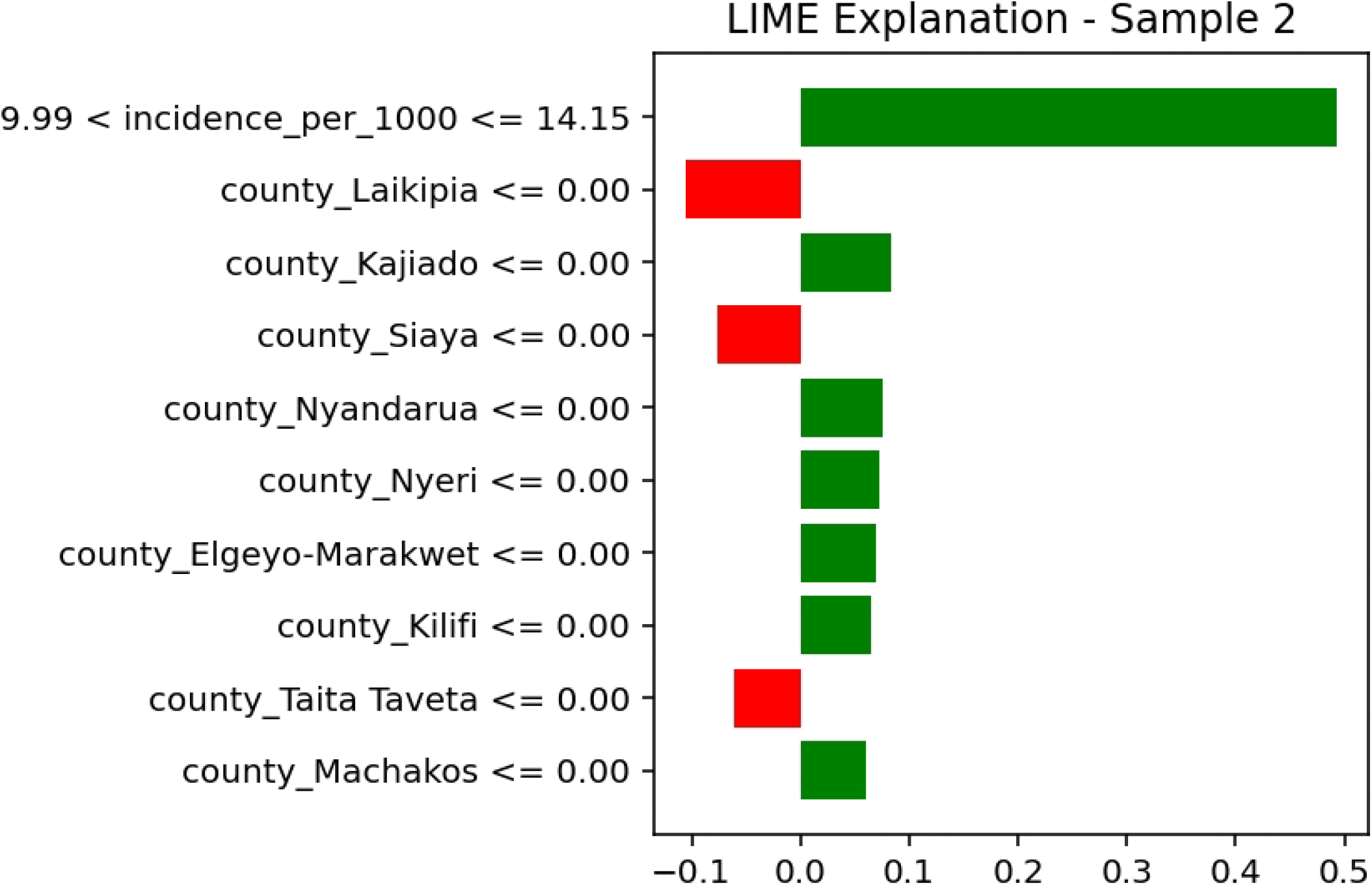
LIME explanation for an instance predicted as class 2 (high transmission). This high-transmission instance is strongly influenced by incidence per 1,000 values in the interval 9.99 ≤ *x* ≤ 14.15, with a positive contribution of approximately 0.45. Additional positive contributions are observed from counties such as Laikipia (0.05), Kajiado (0.08), and Siaya (0.02). In contrast, Taita Taveta contributes a small negative effect (−0.04), slightly reducing the predicted probability of class 2.

## Discussion

This study developed and evaluated a machine learning framework for classifying malaria transmission states in Kenya using spatio temporal panel data. The results show that ensemble tree based methods, particularly extreme gradient boosting, achieved superior predictive performance, strong probabilistic calibration, and consistent interpretability across multiple evaluation criteria. These findings demonstrate that data driven approaches can provide accurate and reliable tools for malaria surveillance and decision making in endemic settings.

### Model Performance and Predictive Accuracy

Extreme gradient boosting consistently achieved the highest performance across all evaluation metrics. This finding is consistent with previous studies that report strong performance of boosting methods in epidemiological prediction tasks [9, 18]. The superior performance can be attributed to the ability of boosting algorithms to capture nonlinear relationships and complex interactions among climatic, environmental, and intervention related variables.

However, the very high performance observed in this study should be interpreted with caution. Malaria transmission exhibits strong temporal persistence, and the inclusion of lagged predictors, particularly past incidence and transmission states, provides substantial predictive information. As a result, part of the predictive performance reflects the continuity of transmission dynamics rather than purely novel pattern discovery. This highlights the importance of temporal validation and careful interpretation of model outputs in epidemiological applications.

Random forest also demonstrated strong performance, although with slightly lower macro averaged F1 score. This difference may reflect the sequential learning strategy of boosting, which iteratively reduces prediction errors. In contrast, multinomial logistic regression showed lower performance, suggesting limited capacity to represent nonlinear dependencies. The support vector machine exhibited the lowest performance, which may be due to sensitivity to parameter selection and challenges in handling complex temporal structures.

### Importance of Probabilistic Calibration

In addition to predictive accuracy, this study emphasised the importance of probabilistic calibration.

Extreme gradient boosting produced well calibrated probability estimates, as indicated by low Brier scores and calibration curves closely aligned with observed outcomes. In contrast, logistic regression and support vector machine models produced less reliable probability estimates.

Calibration is critical in public health decision making because it determines whether predicted risks correspond to actual observed outcomes. Models that are poorly calibrated may produce misleading confidence in predictions, which can result in inefficient allocation of resources or delayed interventions. The results therefore support the inclusion of calibration assessment as a standard component of model evaluation in malaria prediction studies.

### Interpretation of Key Drivers of Transmission

The explainability analysis provides insight into the drivers of malaria transmission. Lagged incidence emerged as the most influential predictor, confirming the strong temporal dependence of malaria dynamics. Environmental variables such as vegetation index and precipitation were also important, reflecting their role in creating suitable conditions for mosquito breeding and survival [4, 5]. Insecticide treated net coverage was associated with reduced transmission probability, consistent with the protective effect of vector control interventions.

Partial dependence analysis revealed nonlinear relationships between temperature and transmission probability, with increasing risk up to a threshold level followed by stabilisation. This pattern aligns with established biological mechanisms governing mosquito development and parasite replication [7]. Seasonal variation in predicted transmission further reflects rainfall driven dynamics that are characteristic of malaria transmission in East Africa [2, 15].

While these findings are consistent with existing epidemiological knowledge, it is important to note that the interpretations are based on statistical associations rather than causal relationships. Explainability methods provide useful approximations of model behaviour but should not be interpreted as definitive evidence of causality.

### Implications for Malaria Surveillance and Control

The classification of malaria transmission into discrete states offers clear practical advantages for public health planning. Unlike continuous prediction of case counts, state based classification aligns directly with operational categories used in malaria control programs. This facilitates identification of areas requiring intensified intervention, enhanced surveillance, or resource reallocation.

The combination of high predictive performance, reliable probability estimates, and interpretability suggests that the proposed framework can support early warning systems and decision support tools. In particular, calibrated probabilities can assist decision makers in prioritising high risk areas under resource constraints. The relatively low computational cost of the best performing model further supports its feasibility for integration into routine surveillance systems.

### Comparison with Existing Studies

Previous studies have applied machine learning techniques to malaria prediction, focusing primarily on continuous outcomes such as incidence or prevalence [11–13]. In contrast, this study models malaria transmission as discrete operational states, which are more directly aligned with decision making frameworks used in practice.

In addition, this study incorporates calibration assessment and explainability methods within a unified framework. These aspects are often overlooked in existing work, despite their importance for practical implementation. The use of forward chaining validation further distinguishes this study by providing a more realistic evaluation of model performance under temporal dependence, in contrast to random validation approaches that may lead to optimistic estimates [18].

### Limitations

Several limitations should be considered. First, the analysis relies on routinely collected surveillance data, which may be affected by reporting delays, underreporting, and variation in diagnostic practices. These factors can introduce measurement error and influence model performance.

Second, the use of lagged outcome variables, while improving predictive accuracy, may limit the ability of the model to generalise to settings with different transmission dynamics. Future work should evaluate model performance under alternative feature specifications to better understand this dependency.

Third, the classification thresholds used to define transmission states may not fully capture regional heterogeneity. Sensitivity analysis using alternative thresholds could provide further insight into the robustness of the results.

Fourth, not all relevant predictors were available, including entomological indicators, indoor residual spraying coverage, and socioeconomic factors. Inclusion of such variables may improve model performance and enhance understanding of transmission dynamics.

Finally, the study focuses on Kenya, and the extent to which the findings generalise to other regions remains uncertain.

### Future Research

Future research should prioritise external validation using independent datasets from other regions to assess generalisability. Incorporating spatial dependence through spatial modelling approaches may improve representation of geographic transmission patterns.

Integration with mechanistic models could enable scenario analysis under varying environmental and intervention conditions. In addition, the development of operational decision support tools that combine predictions, uncertainty estimates, and interpretability outputs could facilitate adoption by public health practitioners.

Further investigation of advanced temporal modelling approaches, including deep learning methods, may provide additional improvements in capturing complex transmission dynamics.

## Conclusion

This study developed and evaluated a machine learning framework for the classification of malaria transmission states in Kenya using longitudinal surveillance data. The findings show that ensemble tree based methods, particularly extreme gradient boosting, achieve high predictive performance, produce reliable probability estimates, and provide consistent interpretability across multiple evaluation criteria.

The results highlight the importance of temporal information in modelling malaria dynamics. The inclusion of lagged incidence and environmental variables improves predictive capability and reflects the persistence and seasonality of transmission. Key predictors identified by the model, including vegetation index, precipitation, and insecticide treated net coverage, are consistent with established epidemiological evidence, supporting the credibility of the model outputs. The incorporation of probabilistic calibration further strengthens the framework by ensuring that predicted risks are meaningful and suitable for decision making.

Modelling transmission as discrete states provides a practical advantage by aligning predictions with operational categories used in malaria control programs. This approach facilitates more direct translation of model outputs into public health action, including early warning, prioritisation of high risk areas, and efficient allocation of limited resources. In this context, the proposed framework offers a useful tool for strengthening malaria surveillance and supporting evidence based intervention strategies.

However, the strong predictive performance observed in this study is partly influenced by temporal dependence in the data, and caution is required when generalising the results to settings with different transmission patterns. Further validation using independent datasets and alternative feature specifications is necessary to assess robustness and generalisability. In addition, incorporating spatial information, entomological indicators, and health system variables may provide a more comprehensive representation of transmission dynamics.

Future research should focus on external validation, integration with spatial and mechanistic modelling approaches, and development of operational decision support systems that combine prediction, uncertainty, and interpretability. Embedding such frameworks within routine surveillance platforms has the potential to enhance real time monitoring and improve the effectiveness of malaria control efforts.

In conclusion, this study demonstrates that machine learning methods, when combined with calibration assessment and interpretability techniques, can provide accurate, transparent, and operationally relevant tools for modelling malaria transmission in endemic settings.

## Data Availability

The data underlying the results presented in this study are publicly available in Zenodo and can be accessed through the following link https://doi.org/10.5281/zenodo.20051237

## Acknowledgements

The authors acknowledge the use of ChatGPT for generating and refining Python code used in data processing and analysis. All outputs were carefully reviewed, validated, and interpreted by the authors to ensure accuracy and scientific integrity.

## Notes

### Competing Interest Statement

The authors have declared no competing interest.

### Funding Statement

The author(s) received no specific funding for this work.

### Author Declarations

Ethical consideration was not required

